# Could widespread use of antiviral treatment curb the COVID-19 pandemic? A modeling study

**DOI:** 10.1101/2021.11.10.21266139

**Authors:** Laura Matrajt, Elizabeth R. Brown, Myron S. Cohen, Dobromir Dimitrov, Holly Janes

## Abstract

Despite the development of safe and effective vaccines, effective treatments for COVID-19 disease are still urgently needed. Several antiviral drugs have shown to be effective in reducing progression of COVID-19 disease. In the present work, we use an agent-based mathematical model to assess the potential population impact of the use of antiviral treatments in four countries with different demographic structure and current levels of vaccination coverage: Kenya, Mexico, United States (US) and Belgium. We analyzed antiviral effects on reducing hospitalization and death, and potential antiviral effects on reducing transmission. For each country, we varied daily treatment initiation rate (DTIR) and antiviral effect in reducing transmission (AVT). Irrespective of location and AVT, widespread antiviral treatment of symptomatic adult infections (≥20% DTIR) prevented the majority of COVID-19 deaths, and recruiting 6% of all adult symptomatic infections daily reduced mortality by a third in all countries. Furthermore, our model projected that targeting antiviral treatment to the oldest age group (65 years old and older, DTIR of 20%) can prevent over 47% of deaths. Our results suggest that early antiviral treatment (as soon as possible after inception of infection) is needed to mitigate transmission, preventing 50% more infections compared to late treatment (started 3 to 5 days after symptoms onset). Our results highlight the synergistic effect of vaccination and antiviral treatment: as the vaccination rate increases, antivirals have a larger relative impact on population transmission. These results suggest that antiviral treatments can become a strategic tool that, in combination with vaccination, can significantly reduce COVID-19 hospitalizations and deaths and can help control SARS-CoV-2 transmission.

## Introduction

With over 5 million deaths worldwide [1], the COVID-19 pandemic has proven difficult to contain. Despite the development, advent, licensure, and rollout of many safe and effective vaccines [2], controlling SARS-CoV-2 transmission has shown to be elusive for several reasons, including vaccine supply shortages in low- and middle-income countries [3], vaccine hesitancy [4], and the emergence of new variants [5]. Indeed, the Delta and Omicron variants, that emerged in summer and fall of 2021, quickly became the predominant strains and have caused large epidemic outbreaks, even in highly vaccinated regions [1]. Rapidly producing such COVID-19 vaccines has been an amazing scientific endeavor, but effective tools to treat COVID-19 disease are still urgently needed. Monoclonal antibodies, antibody cocktails and antiretroviral treatments have been, and continue to be studied to treat SARS-CoV-2 infection and to prevent progression to severe disease [6]. Several treatments have been found to reduce hospitalizations by 30% to 89% [7–10] when taken within the first five days after developing symptoms. Some of them are approved for early treatment of patients with mild-to moderate COVID-19 who are at high-risk of progression to severe disease while others are approved for hospitalized patients, with one approved as a pre-exposure prophylaxis [11]. Furthermore, most studies have shown that these antiviral treatments significantly reduced the amount of infectious virus in the nasal mucosa of treated individuals [12, 13]. Hence, the advent of effective antiviral drugs raises the possibility that in treating infected individuals we may reduce onward transmission (indirect population benefit) while also protecting the treated person from severe disease (direct benefit). The use of antiviral treatments as an effective means of prevention and epidemic control is not new. During the 2009 influenza A H1N1 pandemic, just a few weeks after the first case of influenza A H1N1 was identified in the US, the US government released 11 million courses of antiviral drugs for influenza (25% of the antiviral supply) from the National Stockpile as a potential tool to control transmission and mitigate disease [14]. Treatment as Prevention is considered a primary method of epidemic control for HIV, as research has demonstrated that earliest detection and treatment suppressing HIV replication stops secondary transmission while having the the greatest effect at the individual level [15–17].

Over the past several months, the availability of antigen tests has expanded considerably, facilitating the early diagnosis of SARS-CoV-2 infection and possible early treatment [18, 19]. The US government has purchased 20 million courses of the antiviral pill paxlovid; these are expected to be delivered in early 2022. Furthermore, a “Test to Treat” initiative was recently announced as part of a new phase in the US government pandemic response [20]. The Medicines Patent Pool and the manufacturers of molnupiravir and paxlovid (Merck and Pfizer respectively) have announced license agreements to facilitate global access for these drugs [21, 22], and Pfizer will donate 4 million courses of paxlovid to UNICEF for use in lower-income countries in the following months [23]. Hence, it is possible that in the next few months antiviral treatments will become widely available globally.

In this work, we use an agent-based mathematical model to evaluate the potential population impact of widespread use of antiviral treatments in reducing hospitalization risk and population-level transmission. We explored the use of antiviral treatments in four different countries (Kenya, Mexico, US and Belgium) with very different demographic composition and vastly different proportions of vaccinated individuals. We showed that the synergistic use of vaccine and antiviral treatments can significantly reduce the burden of COVID-19. Further, our model suggested that targeted use of antiviral treatments can be used to prevent the majority of deaths.

## Results

Briefly, we used COVASIM, a previously developed agent-based model of SARS-CoV-2 spread calibrated to Seattle, WA [24, 25]. Our model simulates a population of 500,000 people interacting through a network over the course of 6 months, where each individual in the population is an agent. Every day, individuals contact others in four possible locations: home, school, work or community. At a given point in time, individuals can be susceptible, infected asymptomatic, pre-symptomatic or symptomatic, and recovered. Symptomatic infected individuals can develop a mild, severe or critical infection after symptoms onset. A fixed age-dependent proportion of infections is assumed to remain asymptomatic. Within each age stratum, asymptomatic infections are assumed 30% less transmissible than symptomatic infections (Table S1). Infectivity is time dependent, being highest around symptom onset and decreasing afterwards. We assumed that 40% of the population has been infected in previous epidemic waves and protected from re-infection over the study duration, and compared these results to scenarios assuming 20 and 60% pre-existing immunity. We simulated populations of equal size in four different countries: Kenya, Mexico, US and Belgium. For each country, the model uses country-specific demographics to inform population structure and household sizes. Because we are interested in investigating population effects of antivirals in different epidemic contexts that may modulate their effects, we explored two main scenarios: first, we assumed deployment would take place under an Omicron-like wave, representing a high transmissible variant for which vaccines might not be very effective at preventing infections. In this scenario, we assumed vaccination coverages in each country as of January 3rd 2022, separating the proportion of the population that received full doses or boosters in US and Belgium. Then, we repeated the analysis assuming antiviral deployment under a Delta-like wave, a less transmissible variant for which vaccines remained moderately effective in preventing infections. Here, we assumed vaccination coverage as of October 12th, 2021. For each scenario, we used different parameters for viral transmission, and vaccine effectiveness (Methods for full details).

We assumed that the antiviral treatment would have two primary effects. First, in line with results from the EPIC-HR (paxlovid), PINETREE (remdesivir), COMET-ICE (sotrovimab), and MOVe-OUT (molnupiravir) clinical trials, that showed a very high reduction in hospitalization or deaths, we assumed an antiviral effect on hospitalization (denoted by AVH) by which a course of antiviral treatment would reduce the rate of hospitalization of symptomatic infected individuals by 88% (main results) or 30% (sensitivity analysis) [7–10]. Second, because clinical trials have reported an antiviral effect on viral load, [7, 12, 13], we assumed that the antiviral treatment would have an effect in overall transmission (denoted by AVT). Given uncertainty in how a reduction in viral load translates into a reduction in transmission, we explored reductions in secondary transmission 25, 50, 75, or 100% due to antiviral treatment.

We assumed that antiviral treatment would be given to symptomatically infected adults (18 years or older) within five days from symptoms onset. We explored scenarios with treatment initiated within five days, within two days or between days 3 and 5 from symptoms onset, to study the impact of treatment timing on the results. To reflect real-world constraints on resources and infrastructure, we also varied the rate at which symptomatic adults could be identified and recruited for treatment, considering scenarios with a daily treatment initiation rate (DTIR) of 2, 4, 6, 8, 10, 20, … 100% of symptomatic adults. For example, a 6% DTIR resulted on treating roughly 26% of the overall adult symptomatic infections (total symptomatic infections over the duration of the study), while a DTIR of 40% resulted in treating over 90% of the overall adult symptomatic infections. To capture variability, 100 simulations were run for each scenario. Throughout the text, we present the median percentage deaths and infections averted compared to base-case scenario assuming that existing non-pharmaceutical measures remain in place for the next 6 months without additional vaccination. Cumulative deaths and infections (and respective confidence bounds) are presented in the Supplemental Material.

### Omicron-like wave

#### Targeted use of antiviral treatment can avert large numbers of deaths

We first examine the use of antiviral treatment in all adults. For all the countries considered, irrespective of the antiviral effect on viral transmission, large numbers of deaths were averted for all scenarios, even if the DTIR was low. A 6% DTIR among all adult symptomatic infections averted over 20% of deaths when compared to no antiviral use in all countries (24, 23, 23 and 24% deaths averted for Kenya, Mexico, US and Belgium respectively assuming AVT=25%). Over 58% of deaths would be averted if 20% of all adult symptomatic infections were identified and recruited for treatment daily (58, 58, 59 and 59% of deaths averted for Kenya, Mexico, US and Belgium respectively assuming AVT=25%). If the DTIR increased to 50% or more, over 85% of deaths would be averted in all countries. We observed minimal differences between countries and assumed levels of antiviral reduction in transmission, pointing to the fact that these reductions are a result of direct rather than indirect antiviral treatment protection (Figs. 1 and S1).

**Figure 1:**
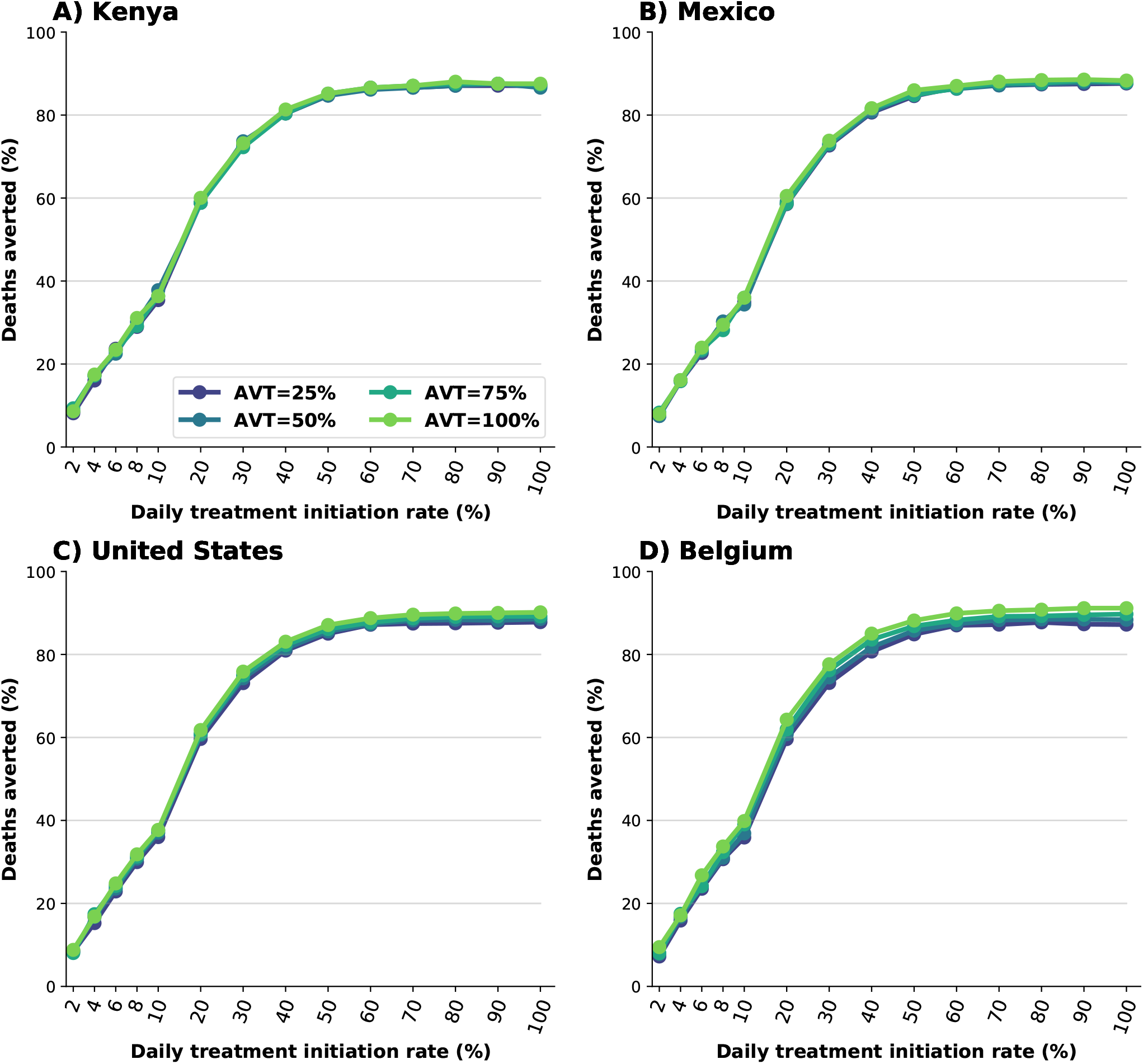
Percentage of deaths averted (compared to a baseline of no antiviral treatment) for A) Kenya, B) Mexico, C) United States and D) Belgium. Here, we assumed an epidemic wave with parameters similar to those of the Omicron epidemic wave (transmissibility, vaccine effectiveness, and vaccination coverage). For each country, the colors represent four possible values of AVT (25, 50, 75 or 100% reduction in viral transmission in treated symptomatic individuals) and a daily treatment initiation rate (DTIR) of 2-100% of adult symptomatic individuals within the first 5 days of symptoms.

Because identifying, recruiting and treating large numbers of all the eligible symptomatic adults (individuals aged 18 or older) might be difficult to implement in practice, especially in countries where testing is not widely available or where antiviral treatment is in short supply, we then evaluated additional targeted strategies where we restricted antiviral treatment to adults over 30, 50 or 65 years of age and considered a 20% DTIR among these populations. Concentrating antiviral treatment in the older age groups can achieve high population impact: despite the fact that the older age groups are the smallest and have the highest vaccination rates (table S4), identifying and treating 20% of the daily symptomatic infections among older adults (aged 65 years old and older) averted 32, 43, 47 and 47% of all deaths in Kenya, Mexico, US and Belgium respectively (assuming 25% reduction in viral transmission). Adults aged 50 and older represent 26, 40 and 44% of the adult populations in Mexico, US and Belgium respectively, but recruiting 20% of the daily symptomatic infections in this age group averted the majority of deaths in these countries (53, 54 and 56% deaths averted in Mexico, US and Belgium respectively). More impressively, while this age group represents only 13% of the adult population in Kenya, initiating antiviral treatment in 20% of the symptomatic infections in this age group daily resulted in 47% of deaths averted. Extending the use of antiviral treatment to younger adults resulted in a marginal gain in Mexico, US and Belgium (a maximum 7, 4 and 6% more deaths averted respectively) when compared with targeted treatment in adults older than 50 (Fig. 2).

**Figure 2:**
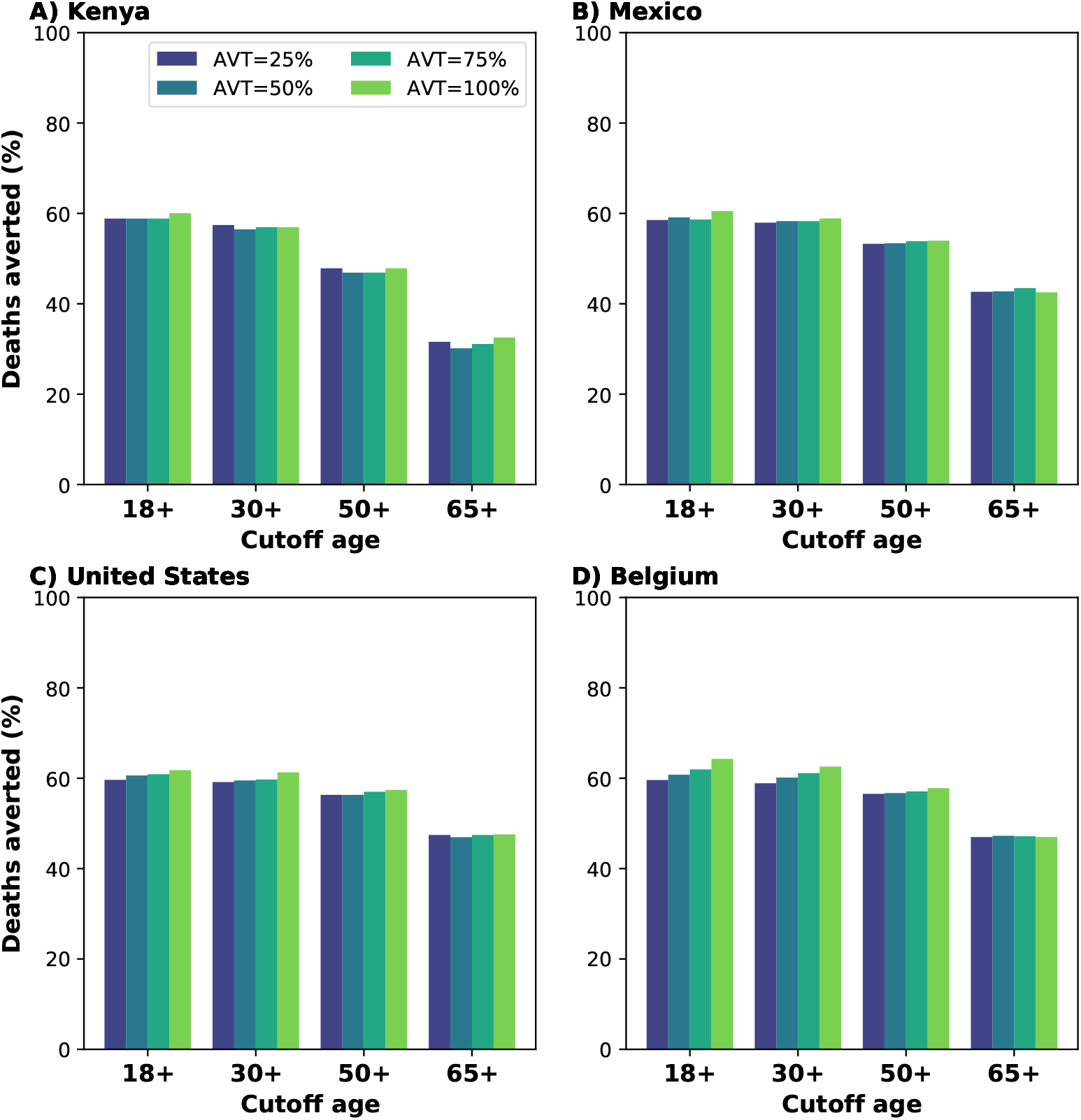
Percentage of deaths averted (compared to a baseline of no antiviral treatment) for A) Kenya, B) Mexico, C) United States and D) Belgium. Here, we assumed an epidemic wave with parameters similar to those of the Omicron epidemic wave (transmissibility, vaccine effectiveness, and vaccination coverage). For each country, the colors represent four possible values of AVT (25, 50, 75 or 100% reduction in viral transmission in treated symptomatic individuals) and targeting the antiviral treatment to symptomatic adults older than 18, 30, 50 or 65 years of age with a daily treatment initiation rate of 20%.

#### Synergistic effect of antiviral treatment and vaccine can mitigate SARS-CoV-2 transmission

We evaluated the potential effect of antiviral treatment on population incidence of SARS-CoV-2, mediated through reduced transmission. Our model projected that antiviral treatment will have a very limited impact in mitigating transmission in the presence of a variant as transmissible as Omicron in countries like Kenya or Mexico that have low vaccination and boosting rates. In highly vaccinated countries like the US and Belgium, our results showed that antiviral treatment had some impact provided that a large proportion of symptomatic infections are identified and recruited for treatment and the antiviral effect on transmission is high (Figs. 3 and S2). This is because the Omicron variant was very transmissible and vaccination with two doses had a minimal effect against preventing infection, but adding a third dose of vaccine provided some protection against acquisition. Further, we found a clear synergy of combining antiviral treatment and vaccination. While Belgium and the US have similar population structure, vaccination coverage in Belgium has been much higher than in the US (both with full doses and with boosters, tables S5 and S6). When combined with antiviral treatment, this difference had a big effect in mitigating transmission: for example, in the most optimistic scenario, assuming that the antiviral treatment completely blocked transmission, a 30% DTIR of adult symptomatic infections averted 9% of overall infections in the US, and twice as many infections were averted in Belgium (18%). In contrast, for this scenario, only 7% of overall infections were averted in Mexico, which had a high vaccination coverage but had not been boosted the population at that time, and 5% of the overall infections were averted in Kenya. Indeed with very limited vaccine supply, the effective reproductive number in Kenya is much larger than in the other countries, resulting in epidemics that are more difficult to control (Figs. 3 and 4). As expected, the antiviral effect on viral transmission played a bigger role in controlling SARS-CoV-2 transmission. With a low AVT (25%), a very limited number of infections were averted, irrespective of treatment initiation rate or country, with a maximum of 6% of infections averted, but a maximum of 35% of infections were averted with high AVT (Belgium, Fig. 3D). Note that, for AVT= 25, 50 or 75%, the number of infections averted plateaued for medium and high antiviral treatment coverage. For example, in the US, a DTIR of 50% of adult symptomatic infections with an antiviral treatment with AVT=50% averted 5% of infections, and treating an additional 20% of symptomatic adults per day (70% DTIR) resulted in only 2% more infections averted (Fig. 3).

**Figure 3:**
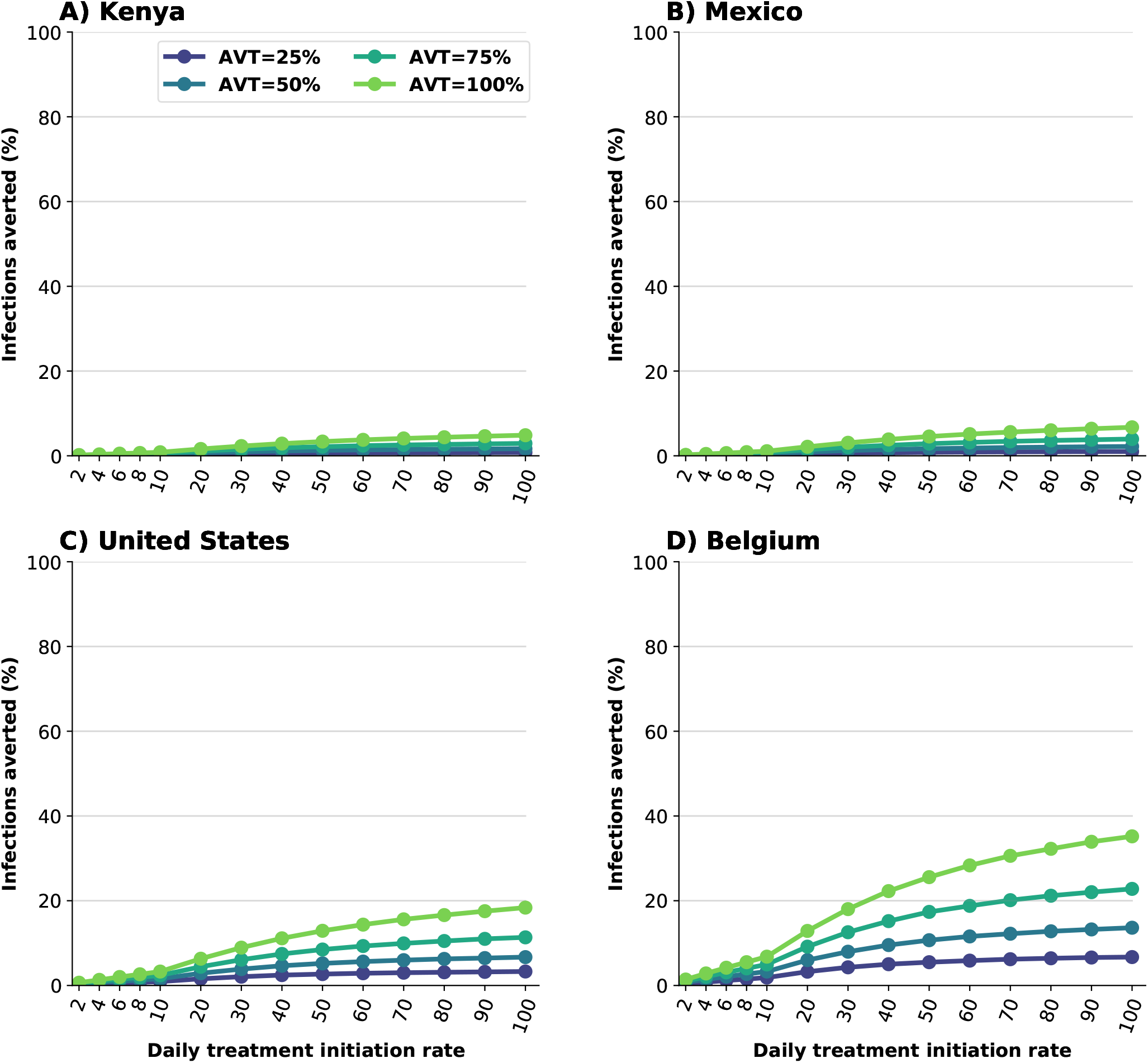
Percentage of infections averted (compared to a baseline of no antiviral treatment) for A) Kenya, B) Mexico, C) United States and D) Belgium. Here, we assumed an epidemic wave with parameters similar to those of the Omicron epidemic wave (transmissibility, vaccine effectiveness, and vaccination coverage). For each country, the colors represent four possible values of AVT (25, 50, 75 or 100% reduction in viral transmission in treated symptomatic individuals) and a daily treatment initiation rate (DTIR) of 2-100% of adult symptomatic individuals within the first 5 days of symptoms.

**Figure 4:**
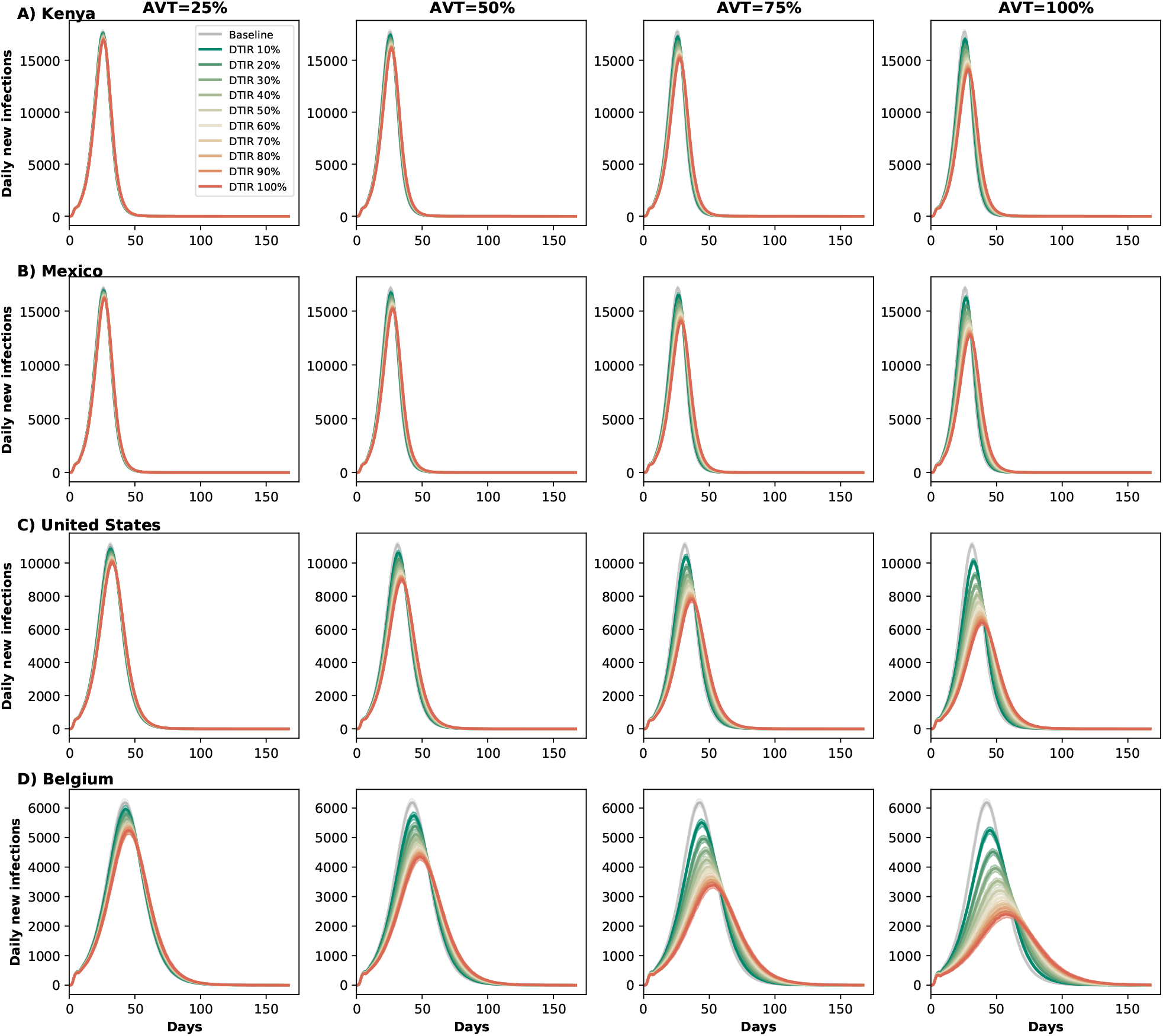
Daily new infections assuming no antiviral treatment (Baseline) or assuming coverage of 10-100% of eligible symptomatic individuals in A) Kenya, B) Mexico, C) United States and D) Belgium. Here, we assumed an epidemic wave with parameters similar to those of the Omicron epidemic wave (transmissibility, vaccine effectiveness, and vaccination coverage). For each location, each column represents a different value of AVT (25, 50, 75 or 100% reduction in viral load).

#### Early treatment is needed to mitigate transmission

We analyzed the difference between early and late treatment by considering treating eligible symptomatic infections within the first two days of symptoms or alternatively between days 3 and 5 after symptom onset. For Kenya and Mexico, late treatment had no impact on transmission (Fig. 5A and B). This is because viral transmission is extremely high and antiviral treatment is happening after most of the secondary infections among symptomatic individuals have occurred. For the US or Belgium, starting antiviral treatment during the first two days of symptom onset prevented roughly twice as many subsequent infections compared to late treatment (Fig. 5C and D). For example, with a DTIR of 50% of symptomatic infections, and AVT = 75%, 14% and 8% of overall infections were averted in Belgium with early and late treatment respectively (Fig. 5D).

**Figure 5:**
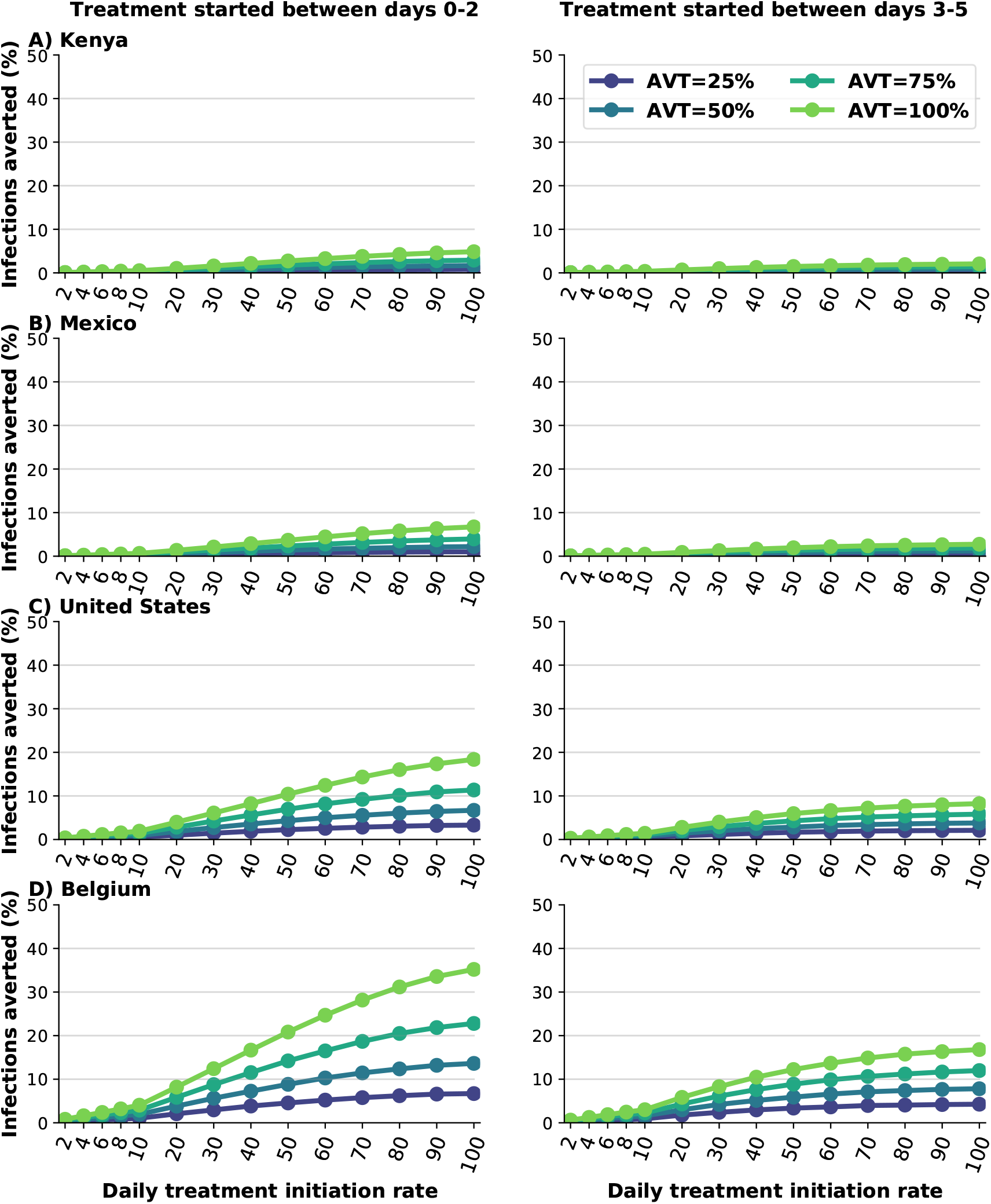
Percentage of deaths averted (compared to a baseline of no antiviral treatment) for A) Kenya, B) Mexico, C) United States and D) Belgium for early (within the first two days of symptoms, left column) or late (between days 2 and 5 of symptoms, right column) treatment. Here, we assumed an epidemic wave with parameters similar to those of the Omicron epidemic wave (transmissibility, vaccine effectiveness, and vaccination coverage). For each country, the colors represent four possible values of AVT (25, 50, 75 or 100% reduction in viral transmission in treated symptomatic individuals) and covering 10-100% of eligible symptomatic individuals.

### Delta-like epidemic wave

In this section we present the results assuming antiviral treatment was given under similar conditions as those experienced during the Delta wave: we considered vaccination coverages as of October 12th, 2021, higher vaccine efficacy (table S2), and a virus as transmissible as the Delta variant (full details in Methods). Under this scenario, the number of deaths averted was slightly higher than for the Omicron wave with an average 2% more deaths averted in all countries (Figs. 1 and S3). Additionally, there were some differences in the number of deaths averted depending on the assumed antiviral effect on transmission, with a maximum of 3, 7, 9 and 12% more deaths averted in Kenya, Mexico, US and Belgium respectively for an antiviral treatment fully blocking transmission compared with one reducing transmission by 25%, suggesting a small indirect effect on the number of deaths averted as a result of reduced transmission. More importantly, under this scenario, we observed that antiviral treatment markedly cut overall viral transmission, and the synergy with vaccination was more apparent. This is because the reduced transmissibility of a Delta-like variant combined with higher vaccine efficacy and higher proportions of the population vaccinated result in a lower effective reproductive number. For example, assuming a DTIR of 30% of symptomatic adults and AVT=50%, 5, 12, 16 and 32% of infections were averted in Kenya, Mexico, US and Belgium respectively (compared to 1, 1, 4 and 8% under an Omicron wave respectively), Figs. 3 and S4.

### Sensitivity analysis

In this section we analyze the sensitivity of our results to key model parameters. All the results presented here assumed an Omicron-type epidemic.

#### Increase or decreased infection-acquired pre-existing immunity

Different places, even within a country, have experienced the COVID-19 pandemic in different ways, depending on local non-pharmaceutical interventions, culture (e.g. vaccine hesitancy, adherence to mask use) and population demographics [26, 27]. Hence, we considered the effect of pre-existing infection-acquired immunity on our results, assuming a lower (20%) or higher (60%) fraction of the population previously infected and currently immune. We observed minimal differences in the number of deaths averted for lower or higher assumed infection-acquired pre-existing immunity. This is expected, as the majority of deaths averted in our model are the result of direct effect of the antiviral treatment. However, the model projected differences in transmission assuming lower or higher infection-acquired pre-existing immunity. Lower infection-acquired immunity resulted in bigger epidemic waves and lower proportions of infections averted with antiviral treatment compared to the main results for all countries, all coverages and all AVEs (maximum 4, 6, 12 and 21% infections averted in Kenya, Mexico, US and Belgium respectively), Figs. S5 left column, S6. In contrast, higher infection-acquired immunity resulted in lower epidemic curves and more infections averted (maximum 13, 18, 41 and 82% infections averted in Kenya, Mexico, US and Belgium respectively), Figs. S5 right column and S7.

#### Reduced antiviral effectiveness against hospitalization, AVH = 30%

One antiviral treatment (molnupiravir) was shown to have a 30% reduction in hospitalizations and deaths [7], so we repeated our analysis assuming this lower value for AVH. As expected, an antiviral treatment which reduces the hospitalization risk of symptomatically infected by 30% averted considerably less deaths than our main scenario (with AVH = 88%). For example, a DTIR of 20% among adult symptomatic infections resulted in over 20% of the deaths averted for all four countries, a ∼40% reduction compared to the main scenario. Under this scenario, there was a stronger synergistic effect of vaccination and with the antiviral effect both on deaths averted and on transmission that was compounded with the antiviral effect on transmission: for example, for a DTIR of 20% and AVT=25% there were 19% and 21% deaths averted in Kenya and Belgium respectively (2% more deaths averted in Belgium), but if AVT=100% there was a 10% difference between the number of deaths averted in Belgium and Kenya (21% and 31% deaths averted in Kenya and Belgium respectively) difference Fig. S8.

#### Decrease in asymptomatic infectiousness

The relative infectivity of asymptomatic individuals remains uncertain [28]. We explored scenarios in which asymptomatic individuals are 50% less infectious than symptomatic individuals. The overall antiviral effect on mortality was not sensitive to this parameter: our model projected nearly identical deaths averted for each scenario considered, Fig. S9. However, the number of infections averted assuming a lower infectivity of asymptomatic infections was higher, with higher differences observed in countries with higher vaccination coverages. For example, under this scenario, 20% of the infections were averted in the US assuming a DTIR of 50% and an antiviral treatment fully reducing transmission, compared with 12% infections averted in the main scenario, Fig. S10.

## Discussion

Despite the existence of effective vaccines, the number of deaths due to COVID-19 globally is still above 7,000 per day, with over 1,000 deaths per day in the US [1], due to issues with supply, hesitancy and logistics. This highlights the need of effective and affordable treatment options. Several antiviral treatments have shown to be highly efficacious against COVID-19 hospitalization and death, provided that treatment is started early, within 5 days of symptoms onset [7–10]. In the present work we analyzed the potential population impact of antiviral treatments for reducing SARS-CoV-2 transmission and COVID-19 related hospitalizations and deaths. We considered four equal-sized populations from four different countries (Kenya, Mexico, US and Belgium), representing different population structures and vaccination coverage. We further explored the impact of this intervention under epidemic waves with parameters similar to the Delta and Omicron waves. Our results suggest that irrespective of country, AVT and variant, widespread use of antiviral treatment (daily treatment initiation rate of 20% of the daily adult symptomatic infections) could prevent the majority of deaths. We projected that under an Omicron-like epidemic wave, with extremely high viral transmissibility and low vaccine effectiveness against infection acquisition, antiviral treatment of symptomatic infections will have very limited impact on transmission. This is expected as a large proportion of the infection chain is occurring in asymptomatic infections, and treating symptomatic infections under high transmissibility is not sufficient to curb overall transmission. Under this scenario, expanding antiviral treatment access to all detected infections might increase the effectiveness of antiviral treatments in this regard. However, if newer variants behaved more like the Delta variant, then our model showed that antiviral treatment in symptomatic adults can have a larger impact on curbing overall infections. Considerable practical challenges would be faced in identifying, testing, and treating symptomatic infections among all adults, especially among the younger adults, who are at much lower risk to progress to severe disease and death.

A clinical outcome of considerable interest is long COVID, to which all age groups appear to be susceptible [29, 30]. Long COVID consists of persistent symptoms including abnormal breathing, headache, fatigue, muscle weakness, anxiety or depression, headache, myalgia, and cognitive dysfunction [31]. To date, it is not known if, how and which (if any) antiviral treatments are effective against any of these post-COVID conditions. If antiviral treatments (either monoclonal antibodies or antiviral chemotherapy) were shown to be effective against long COVID, this could justify widespread use of antiviral treatments among all strata of the population. Of course, continuing promoting and financing wide scale testing would be critical for achieving widespread antiviral use.

Moreover, our model suggests that there is a synergistic effect of combining antiviral treatment and vaccination: countries with larger proportion of their populations vaccinated are expected to benefit more from adding antiviral treatment to their pandemic control toolbox. This emphasizes the need of continuous effort to improve vaccination coverage especially in settings where it is extremely low. Finally, our simulations showed that early treatment, when viral transmission is highest [32, 33], is important for mitigating transmission and could be used as a prevention tool. This is in agreement with previous work [34–38] that has shown the potential use of antiviral therapies to reduce COVID-19 related mortality.

Our model, like all mathematical models, is subject to several limitations. We did not consider the development of antiviral resistance, yet it is possible that if antiviral treatments are widely used, resistance could rapidly develop. In fact, monoclonal antibody treatments that were highly effective against older variants have become ineffective against newer ones [39, 40]. However, small molecule oral antiviral treatments have been shown to remain efficacious against new variants [41]. We analyzed the use of antiviral treatment in symptomatic individuals, and did not explore its use in asymptomatic infections or as a prophylaxis. Further, we assumed that an antiviral effect reducing transmission was independent of when treatment was started (as long as it started within the first five days of treatment). In reality, it is possible that antiviral treatments might have different effects in reducing overall transmission depending on when during the course of an infection they are taken, e.g. reducing overall viral load and hence transmission if taken early on but having only a modest effect as time goes by. Because studies have shown contradictory results regarding the effect of vaccines in reducing infectiousness [42–45], we conservatively assumed that the vaccine had no effect in reducing infectiousness. If vaccination does reduce infectiousness then the synergy between antiviral and vaccine might be less than reported here. We assumed no further vaccination during the period of our analysis, which corresponds to the current situation in most middle- and high-vaccinated countries, where the adult population who are willing to be vaccinated and boosted has mostly been immunized. Nevertheless, vaccines for children younger than 5 years old are currently in clinical trials, and are expected to be available during summer 2022. Vaccinating this age group will increase the overall proportion of the population vaccinated in high-income countries, hence reducing further on-going transmission. In addition, WHO recently released an ambitious plan to vaccinate 70% of the global population by mid 2022 [46]. Because we modeled four different countries with different non-pharmaceutical interventions and different cultures, we did not model any additional interventions, such as masking, or behavioral changes among diagnosed people. For simplicity, we did not include waning immunity, but explored scenarios where a higher or lower proportions of the population are currently immune. While it is extremely hard to predict when and how different populations will become vaccinated and the percentage of pre-existing immunity in each population, we believe that the scenarios that we have considered here, with different combinations of vaccination coverage and proportions of the population with pre-existing infection-induced immunity, are sufficient to capture the potential population-level effect of the potential use of antiviral treatment on transmission and COVID-19-related severe outcomes.

Taken together, our results suggest that antiviral treatment can be an extremely useful tool to reduce COVID-19 related deaths and to alleviate the COVID-19 burden on overwhelmed and exhausted healthcare systems. In particular, antiviral treatments, especially oral antiviral drugs that can be easily distributed, can have a huge impact in countries which have had less access to vaccines and boosters. Further, our model shows that the population level impact of antiviral treatments are enhanced by their synergistic use in combination with vaccination, particularly in the presence of less transmissible variants. Our results suggest that in the face of highly transmissible variants, unless antiviral treatments are shown to protect against or diminish long COVID conditions, antiviral treatment is best used by targeting it to groups being at high risk for disease progression. As more data emerges, mathematical modeling can be extremely useful to determine the optimal use of antiviral treatments.

## Methods

### Main model

We adapted a previously developed agent-based transmission model, COVASIM [24]. Namely, we extended the model to include the use of antiviral treatments in the population. We briefly describe here the main features of this model (as were used in the present work) and we describe in full detail the adaptations we made. We refer to the original article by Kerr et al. [24] for the full details of the model implementation. This is an agent-based model that simulates SARS-CoV-2 transmission and interventions that was originally calibrated to data for Seattle, WA, US. Each individual in the population is modeled as an agent in a network, with 500,000 agents. The model has four possible contact layers: home, school, work or community. For each location, the model uses country-specific demographics and household sizes. The model simulates infections, interventions antiviral treatment and vaccination. We did 100 runs for each scenario. We report the median of these runs and the 10th and 90th percentiles for the lower and upper bounds.

### Disease dynamics

Individuals in the network can be susceptible, exposed (infected but not infectious), asymptomatic, presymptomatic or symptomatic. Symptomatic individuals have one of three fates: they develop a mild, severe, or critical disease. All mild infected individuals recover, and infected individuals reaching a critical state can recover or die. The latent period is sampled from a log-normal distributions with a mean of 4.6 days. The length of time between becoming infectious and developing symptoms is sampled from a log-normal distribution with mean 1.1 days. The times to develop severe symptoms, to progress to become critically ill and to death are sampled from lognormal distributions with means 6.6, 1.5 and 10.7 days respectively. Asymptomatic and mild cases recover on average on 8 days (sampled from log-normal distributions), while severe and critically ill cases recover on average on 18.1 days. Infectiousness was assumed to be linearly correlated to viral load [47]. As with the original model [24], we modeled viral load having two modes: first, a high mode where viral load is highest, around or before symptom onset, and a low mode having a longer duration but a lower viral load (50% lower than during the high mode). We assumed that children are less likely to develop symptoms than adults but equally likely to become infected [48, 49], Table S3. Further, asymptomatic individuals are assumed to be 30% less infectious than symptomatic individuals [50].

### Viral transmission

We assumed that antiviral treatment would be deployed under epidemic waves with characteristics similar to the Delta or the Omicron waves. For the Delta wave, we assumed an increased transmissibility of 97% with respect to the ancestral variant [5]. For the Omicron wave, we assumed an increased transmissibility of 66% with respect to the Delta variant [51]. These assumptions resulted in basic reproduction number *R*_0_ for a Delta-like variant of approximately 4.95-5.68 and of 7.75-8.63 for an Omicron-like variant. We assumed that 0.5% of the population is infected at the beginning of the simulation and that 40% of the population (selected at random) in each country was previously infected and is immune for the duration of the study (20 and 60% were also explored in Sensitivity analysis). Table S1 summarizes all the parameters used in the model.

### Antiviral treatment

As described above, we assumed that antiviral treatment had two main effects. First, antiviral treatment reduced the probability of symptomatic infected individuals becoming hospitalized. Second, we reduced viral load in treated individuals by 25, 50, 75 or 100%. Because in our model, viral load linearly correlates with infectiousness, this assumption resulted in equal reductions in the overall transmissibility of treated infected individuals. Each day of the simulation, the model identified eligible individuals (symptomatic adults over 18, 30, 50 or 65 years of age) whose symptoms onset was within the time frame studied (5 days, 2 days or within 3-5 days of symptoms onset). It then randomly selected a percentage of these individuals (excluding those who are already in treatment) for antiviral treatment initiation. Once an infected symptomatic individual initiated treatment, we assumed that antiviral effects would last for the subsequent days of his or her infection.

### Vaccination

We considered a leaky vaccine (that is, a vaccine that confers partial protection to all vaccinated individuals) [52] having three effects on vaccinated individuals: to reduce the probability of acquiring a SARS-CoV-2 infection (denoted byVE_SUS_), to reduce the probability of developing COVID-19 symptoms after infection (denoted by VE_SYMP_), and to reduce the probability of hospitalization conditioned on symptomatic infection (denoted by VE_H_). Then it follows that

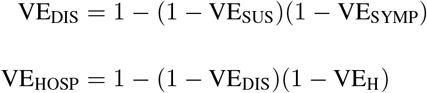

where VE_DIS_ and VE_HOSP_ are the unconditional vaccine effectiveness on symptomatic infection and hospitalization respectively. Importantly, under this model vaccination does not influence onward transmission of infection except through a reduction in SARS-CoV-2 infection. Vaccine effectiveness estimates for each modeled wave can be found in table S2. For Mexico, the US and Belgium, we used country- and age-specific vaccination rates (vaccination rates as of October 12th 2021 and January 3rd, 2022 for the Delta and Omicron waves respectively), tables S5-S7 [53–56]. For the US and Belgium, vaccinated individuals were further split between those having high (boosted individuals) or low protection for the Omicron wave. Since Kenya had fully vaccinated only a very small proportion of its population –1.5% and 7.7% of its population for each of these dates– and targeted front-line health workers, teachers, police and military, we distributed its vaccine randomly among all adults [3, 56, 57].

## Data Availability

Code will be available upon publication.

https://github.com/lulelita/antivirals_covid19

## Author contributions

LM, ERB and HJ conceived the study. LM conducted the analysis and wrote the first draft of the manuscript. HJ, DD, and ERB contributed to writing the manuscript.

## Code availability

Code will be available at: https://github.com/lulelita

## Supplemental Information

**Table S1:** Parameters used in the model.

| Parameter | Distribution (mean, SD) | Reference |
| --- | --- | --- |
| Latent period | lognormal(4.5, 1.5) | [24, 58] |
| Infectious period for asymptomatic and mild cases | lognormal(8, 2) | [24, 59] |
| Duration of presymptomatic period | lognormal(1.1, 0.9) | [24, 60] |
| Length of time from symptom onset to hospitalization | lognormal(6.6, 4.9) | [24, 60, 61] |
| Length of time from hospitalization to critically ill | lognormal(1.5, 1) | [24, 61, 62] |
| Length of time from critically ill to death | lognormal(10.7, 4.8) | [24, 63] |
| Time from onset of symptoms to recovery for severe and critically ill cases | lognormal(18.1, 6.3) | [24, 63] |
| Age-stratified mortality rates | varied by age | [24, 64, 65] |
| Age-stratified probability of developing symptoms | Table S3 | [24, 63, 66] |
| Fraction of symptomatic infections <15 year old | 0.25 | [67] |
| Fraction of symptomatic infections ≥15 year old | 0.6 | [50, 68, 69] |
| Antiviral effect on viral transmission, AVT | 25, 50, 75, 100 | assumed |
| Antiviral effect on hospitalization, AVH, | 50 or 80 | [70, 71] |

**Table S2:** Vaccine effectiveness values used in the model during the Delta and the Omicron waves. For the Delta wave, vaccine effectiveness assumes full coverage. For the Omicron wave, we assumed that boosted vaccinated individuals will get the same protection as that given by full coverage during the Delta wave.

| Parameter | Delta | Omicron (full coverage) | Omicron (boosted) | References |
| --- | --- | --- | --- | --- |
| Vaccine efficacy against infection, $VE_{SUS}$ | 59.6 | 10 | 59.6 | [72–74] |
| Vaccine efficacy against symptomatic infection, $VE_{DIS}$ | 67.5 | 0.15 | 67.5 | [72, 74] |
| Vaccine efficacy against Hospitalization, $VE_{HOSP}$ | 87.65 | 0.52 | 87.65 | [72, 74] |

**Table S3:** Age-specific parameters for disease progression.

| Parameter | 0-9 | 10-19 | 20-29 | 30-39 | 40-49 | 50-59 | 60-69 | 70-79 | 80-89 | 90+ |
| --- | --- | --- | --- | --- | --- | --- | --- | --- | --- | --- |
| Probability of severe disease | 0.0005 | 0.00165 | 0.0072 | 0.0208 | 0.0343 | 0.0765 | 0.1328 | 0.20655 | 0.2457 | 0.2457 |
| Probability of critical disease | 0.00003 | 0.00008 | 0.00036 | 0.00104 | 0.00216 | 0.00933 | 0.03639 | 0.08923 | 0.1742 | 0.1742 |
| Probability of death | 0.00002 | 0.00002 | 0.0001 | 0.00032 | 0.00098 | 0.00265 | 0.00766 | 0.02439 | 0.08292 | 0.1619 |

**Table S4:** Demographic distribution of the adult population

| Table S4: Demographic distribution of the adult population |  |  |  |  |
| --- | --- | --- | --- | --- |
| Location | Kenya | Mexico | US | Belgium |
| Percentage of adults $\geq 18$ | 100 | 100 | 100 | 100 |
| Percentage of adults $\geq 30$ | 44 | 59 | 70 | 74 |
| Percentage of adults $\geq 50$ | 13 | 26 | 40 | 44 |
| Percentage of adults $\geq 65$ | 3 | 9 | 19 | 22 |

**Table S5:**
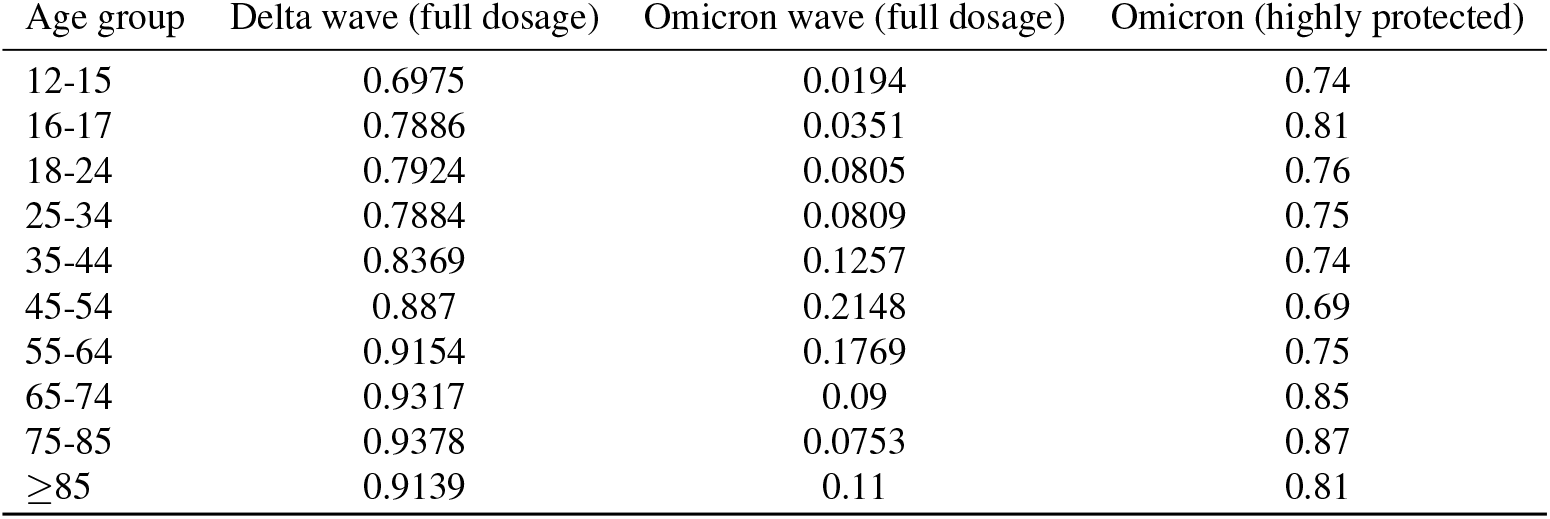
Vaccine distribution in Belgium (values taken from [55, 56]). As of January 3rd, certain groups (e.g. children who just got vaccinated) in Belgium were were considered “highly protected” and were given the vaccine effectiveness of the boosted vaccinated individuals in the model.

| Age group | Delta wave (full dosage) | Omicron wave (full dosage) | Omicron (highly protected) |
| --- | --- | --- | --- |
| 12-15 | 0.6975 | 0.0194 | 0.74 |
| 16-17 | 0.7886 | 0.0351 | 0.81 |
| 18-24 | 0.7924 | 0.0805 | 0.76 |
| 25-34 | 0.7884 | 0.0809 | 0.75 |
| 35-44 | 0.8369 | 0.1257 | 0.74 |
| 45-54 | 0.887 | 0.2148 | 0.69 |
| 55-64 | 0.9154 | 0.1769 | 0.75 |
| 65-74 | 0.9317 | 0.09 | 0.85 |
| 75-85 | 0.9378 | 0.0753 | 0.87 |
| $\geq 85$ | 0.9139 | 0.11 | 0.81 |

**Table S6:**
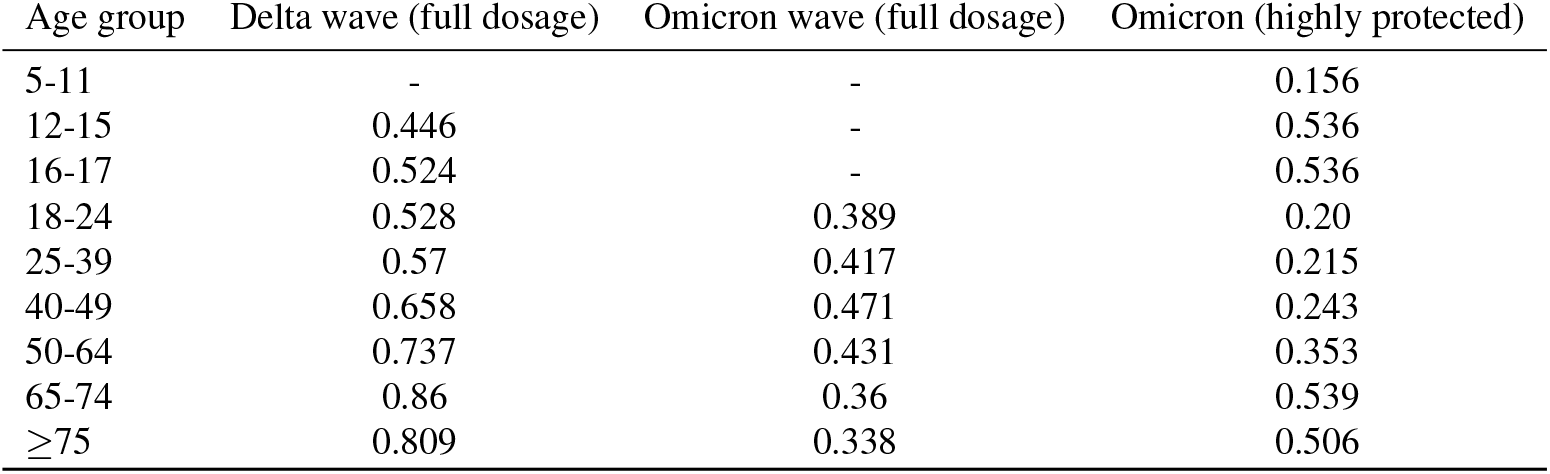
Vaccine distribution in US (values taken from [54, 56]). As of January 3rd, children under 18 years old were considered “highly protected” and were given the vaccine effectiveness of the boosted vaccinated individuals in the model.

**Table S7:** Vaccine distribution in Mexico (values taken from [53, 56]). As of January 3rd, we found no data on boosted individuals in Mexico, so we did not considered boosted individuals in Mexico [56].

| Age group | Delta wave (full dosage) | Omicron wave (full dosage) |
| --- | --- | --- |
| 18-39 | 0.493 | 0.83 |
| 40-49 | 0.735 | 0.735 |
| 50-59 | 0.785 | 0.785 |
| 60 | 0.824 | 0.824 |

**Figure S1:**
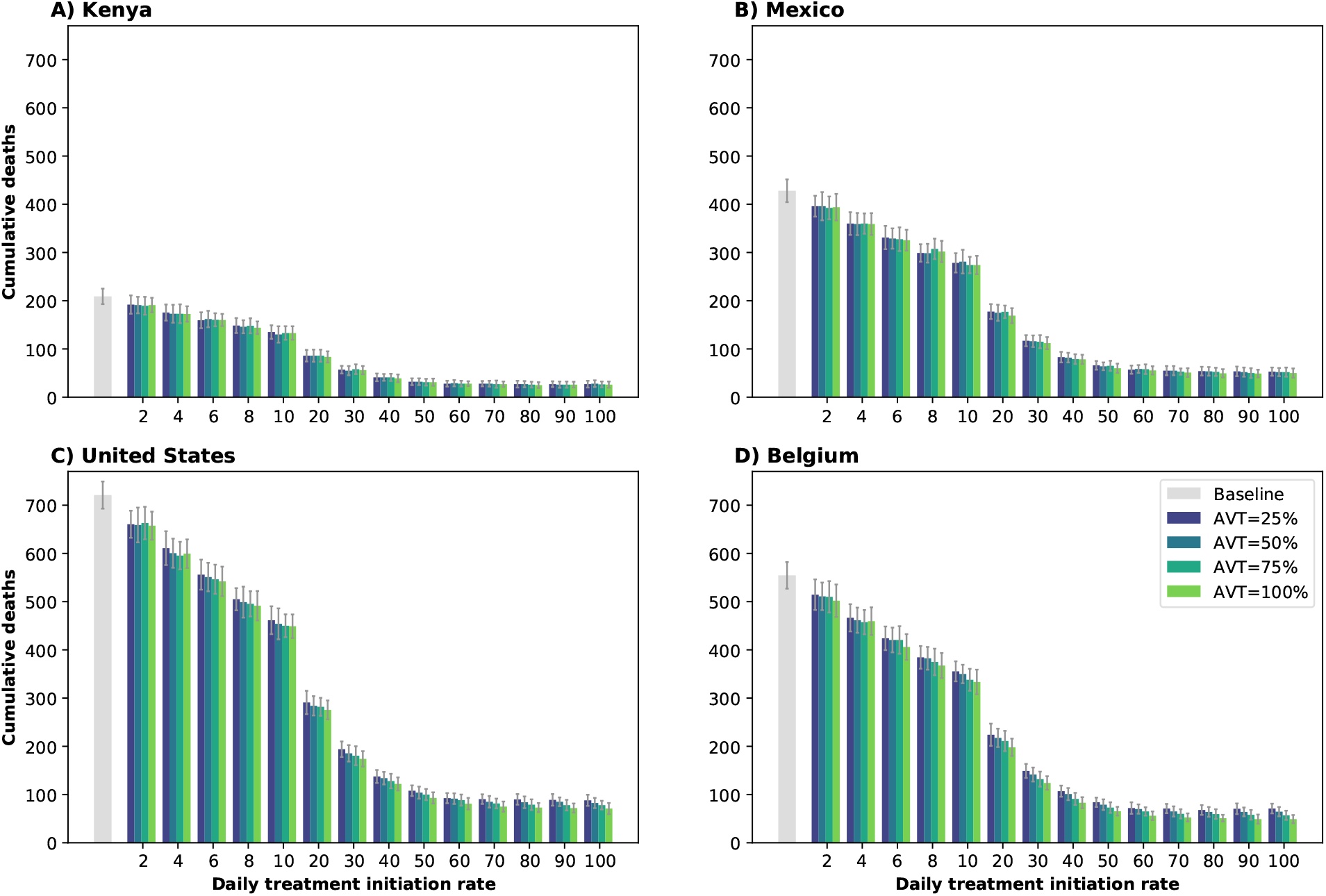
Cumulative deaths over next 6 months in A) Kenya, B) Mexico, C) United States and D) Belgium. Here, we assumed an epidemic wave with parameters similar to those of the Omicron epidemic wave (transmissibility, vaccine effectiveness, and vaccination coverage). For each country, the colors represent four possible values of AVT (25, 50, 75 or 100% reduction in viral transmission in treated symptomatic individuals) and a daily treatment initiation rate (DTIR) of 2-100% of adult symptomatic individuals within the first 5 days of symptoms. Gray bars represent baseline cumulative deaths in absence of antiviral treatment.

**Figure S2:**
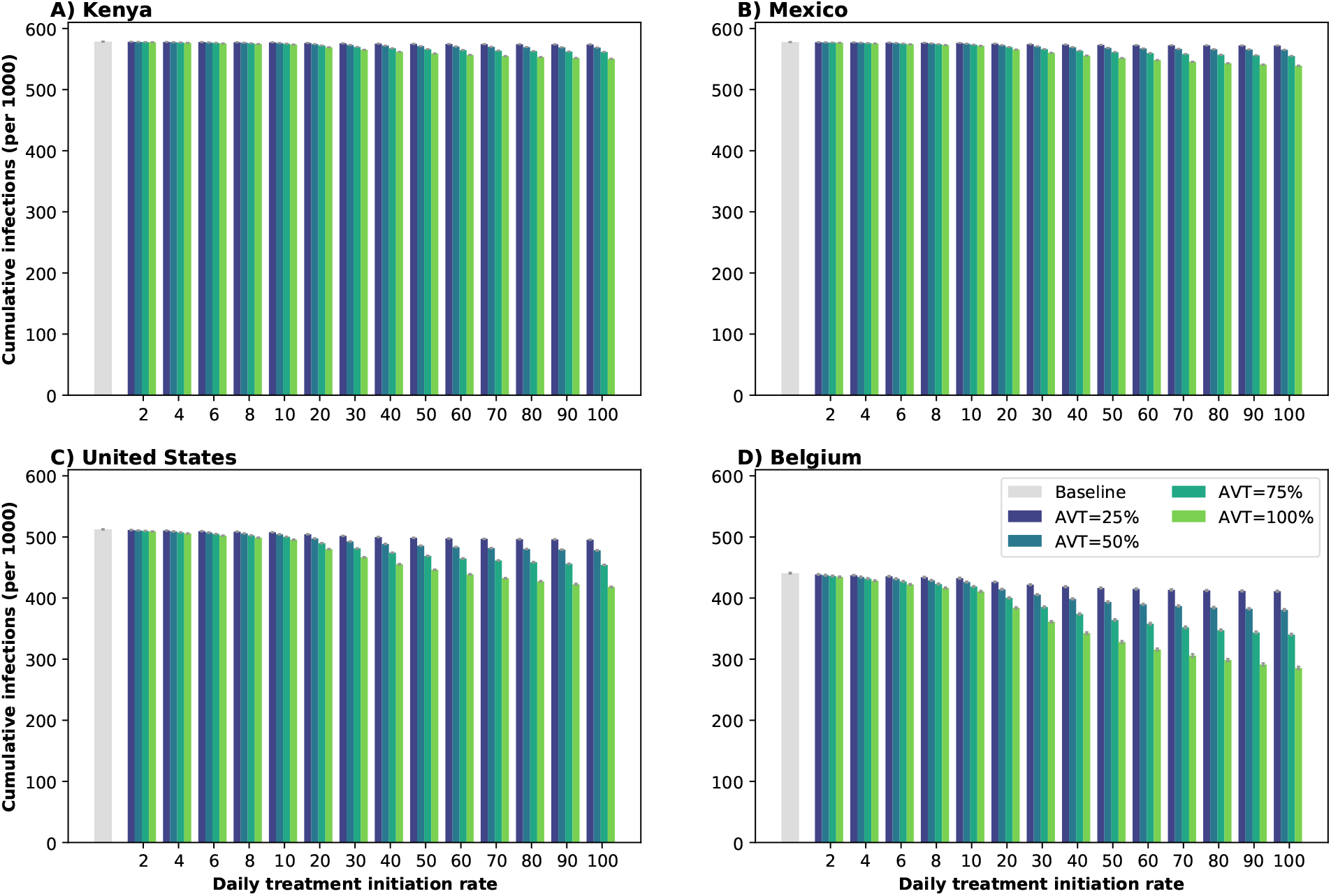
Cumulative infections over next 6 months for A) Kenya, B) Mexico, C) United States and D) Belgium. Here, we assumed an epidemic wave with parameters similar to those of the Omicron epidemic wave (transmissibility, vaccine effectiveness, and vaccination coverage). For each country, the colors represent four possible values of AVT (25, 50, 75 or 100% reduction in viral transmission in treated symptomatic individuals) and a daily treatment initiation rate (DTIR) of 2-100% of adult symptomatic individuals within the first 5 days of symptoms. Gray bars represent baseline cumulative infections in absence of antiviral treatment.

**Figure S3:**
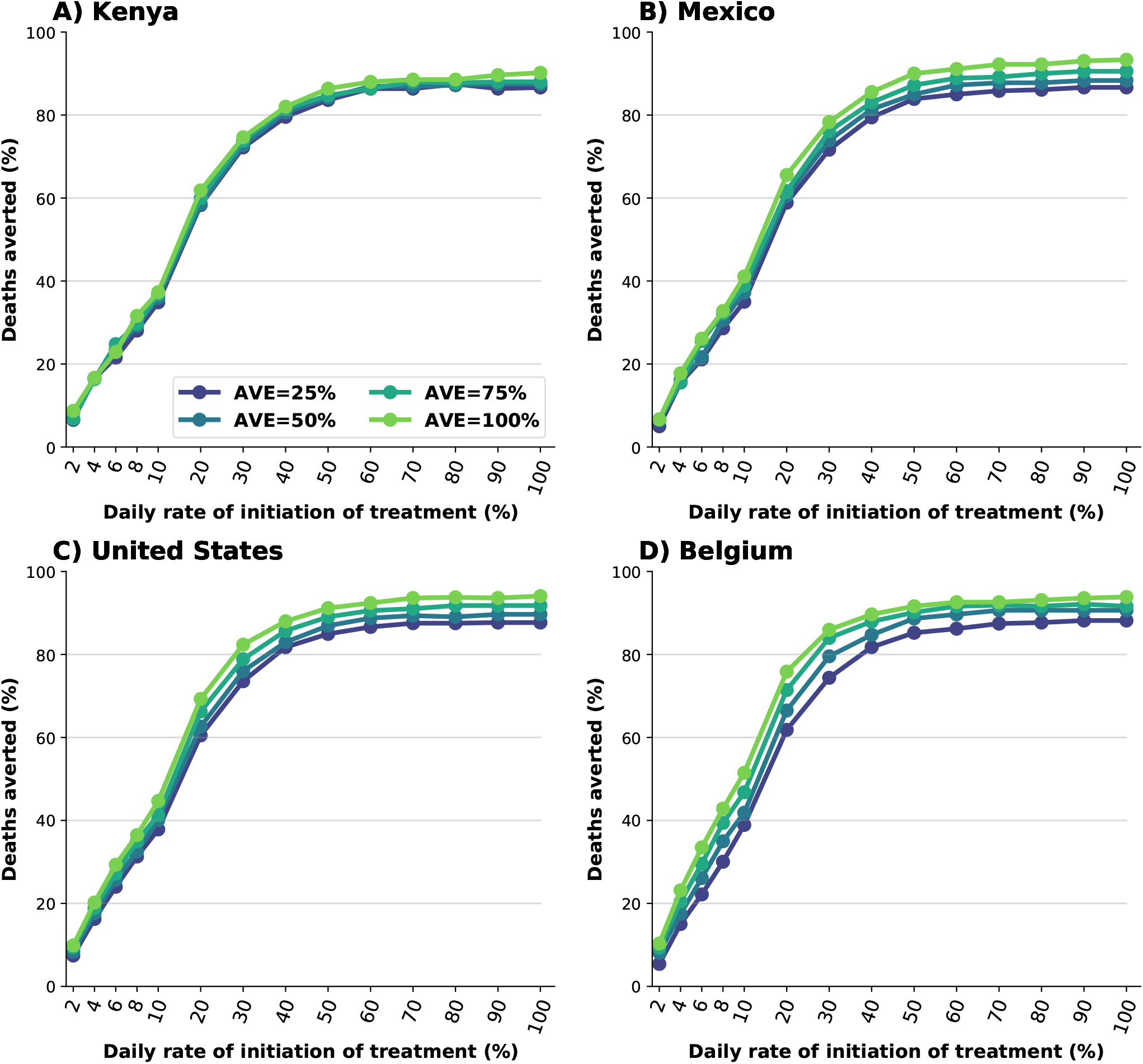
Percentage of deaths averted (compared to a baseline of no antiviral treatment) for A) Kenya, B) Mexico, C) United States and D) Belgium. Here, we assumed an epidemic wave with parameters similar to those of the Delta epidemic wave (transmissibility, vaccine effectiveness, and vaccination coverage). For each country, the colors represent four possible values of AVT (25, 50, 75 or 100% reduction in viral transmission in treated symptomatic individuals) and a daily treatment initiation rate (DTIR) of 2-100% of adult symptomatic individuals within the first 5 days of symptoms.

**Figure S4:**
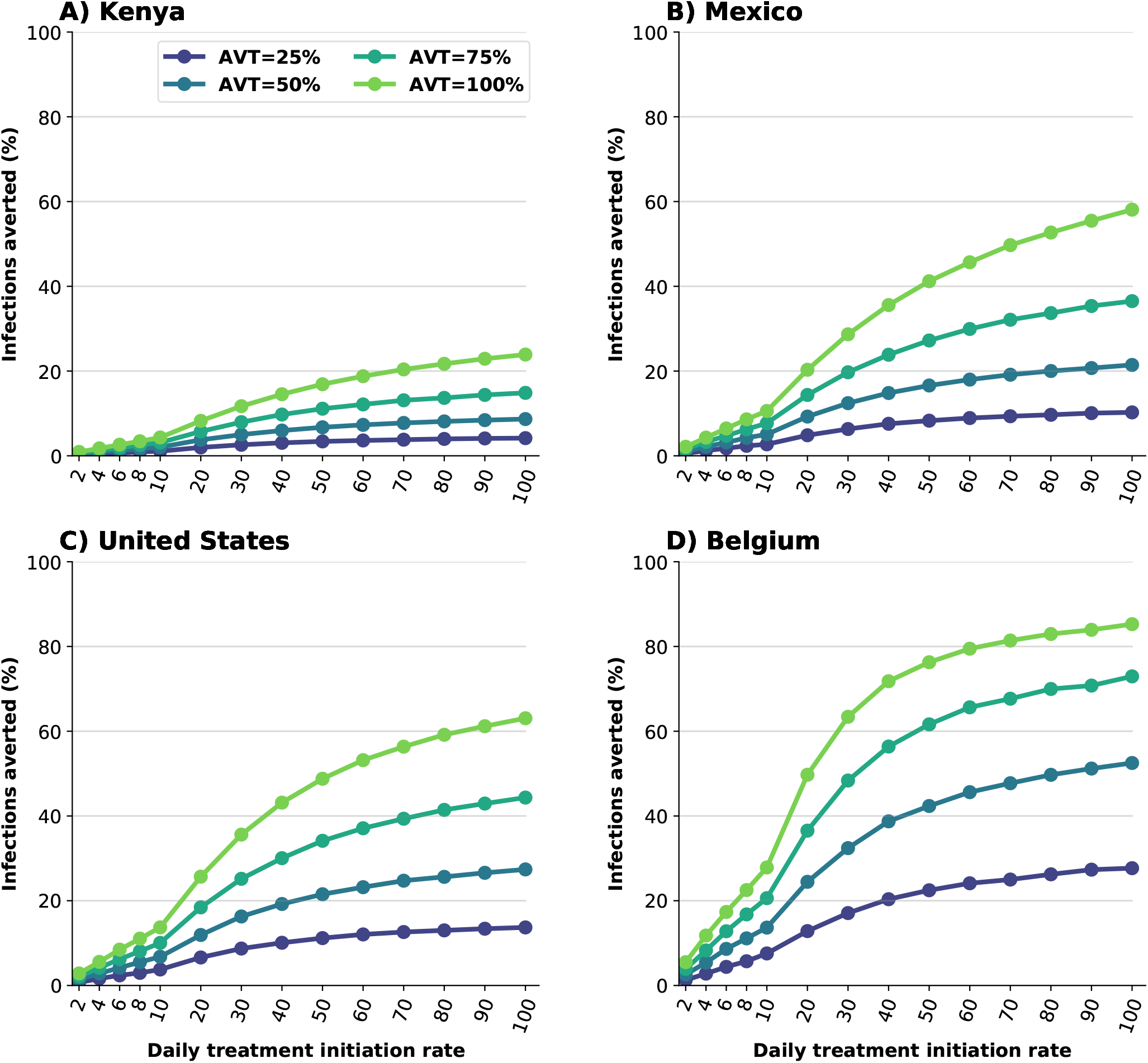
Percentage of infections averted (compared to a baseline of no antiviral treatment) for A) Kenya, B) Mexico, C) United States and D) Belgium. Here, we assumed an epidemic wave with parameters similar to those of the Delta epidemic wave (transmissibility, vaccine effectiveness, and vaccination coverage). For each country, the colors represent four possible values of AVT (25, 50, 75 or 100% reduction in viral transmission in treated symptomatic individuals) and a daily treatment initiation rate (DTIR) of 2-100% of adult symptomatic individuals within the first 5 days of symptoms.

**Figure S5:**
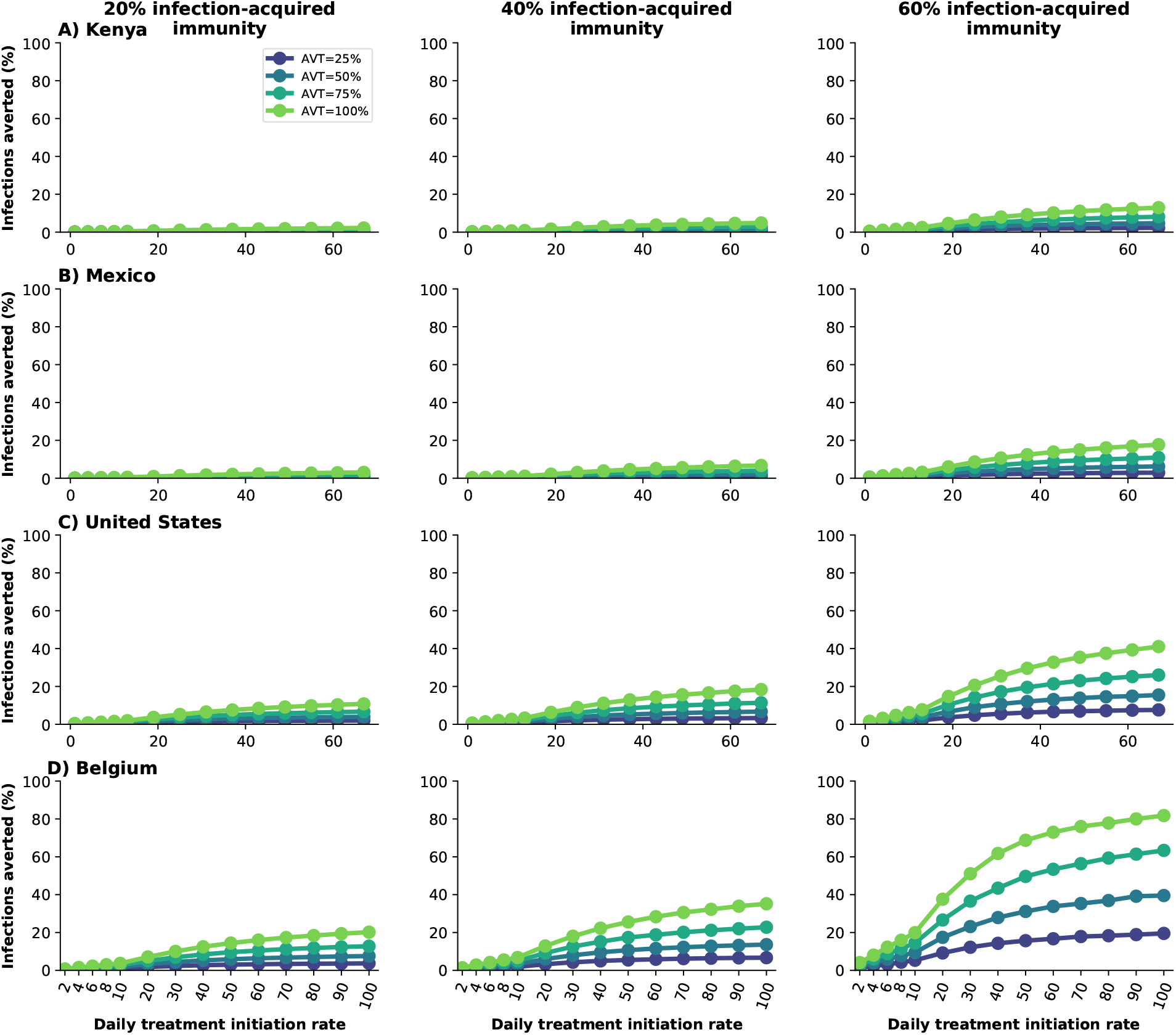
Percentage of deaths averted for A) Kenya, B) Mexico, C) United States and D) Belgium assuming 20 (left), 40 (middle) or 60% (right) of the population has been previously infected and is currently immune. For each country, the colors represent four possible values of AVT (25, 50, 75 or 100% reduction in viral transmission in treated symptomatic individuals) and a daily treatment initiation rate (DTIR) of 2-100% of adult symptomatic individuals within the first 5 days of symptoms.

**Figure S6:**
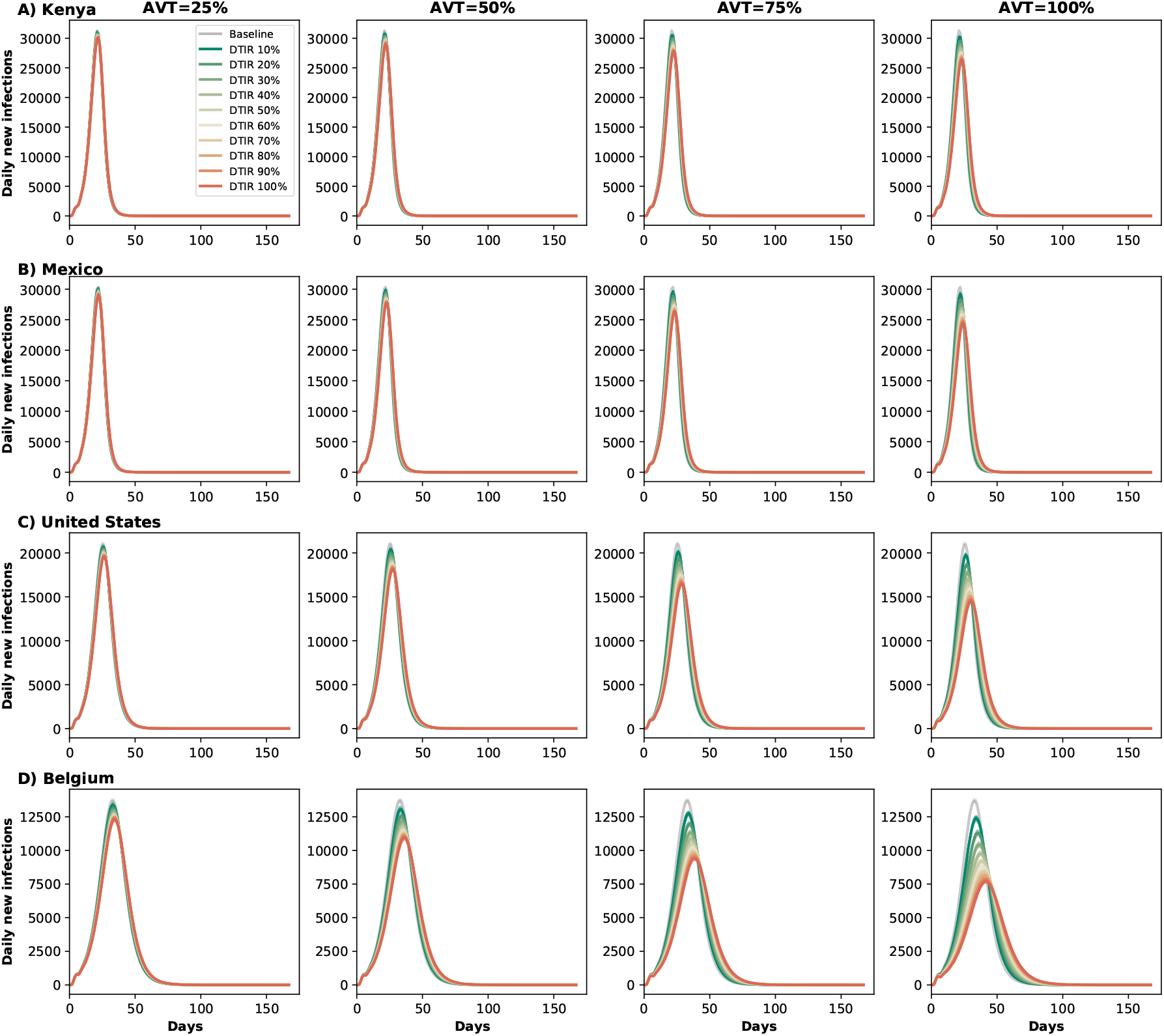
Daily new infections assuming no antiviral treatment (Baseline) or assuming a daily treatment initiation rate of of 10-100% of eligible symptomatic individuals in A) Kenya, B) Mexico, C) United States and D) Belgium assuming 20% of the population has been previously infected and is now recovered. For each location, each column represents a different value of AVT (25, 50, 75 or 100% reduction in viral transmission in treated symptomatic individuals).

**Figure S7:**
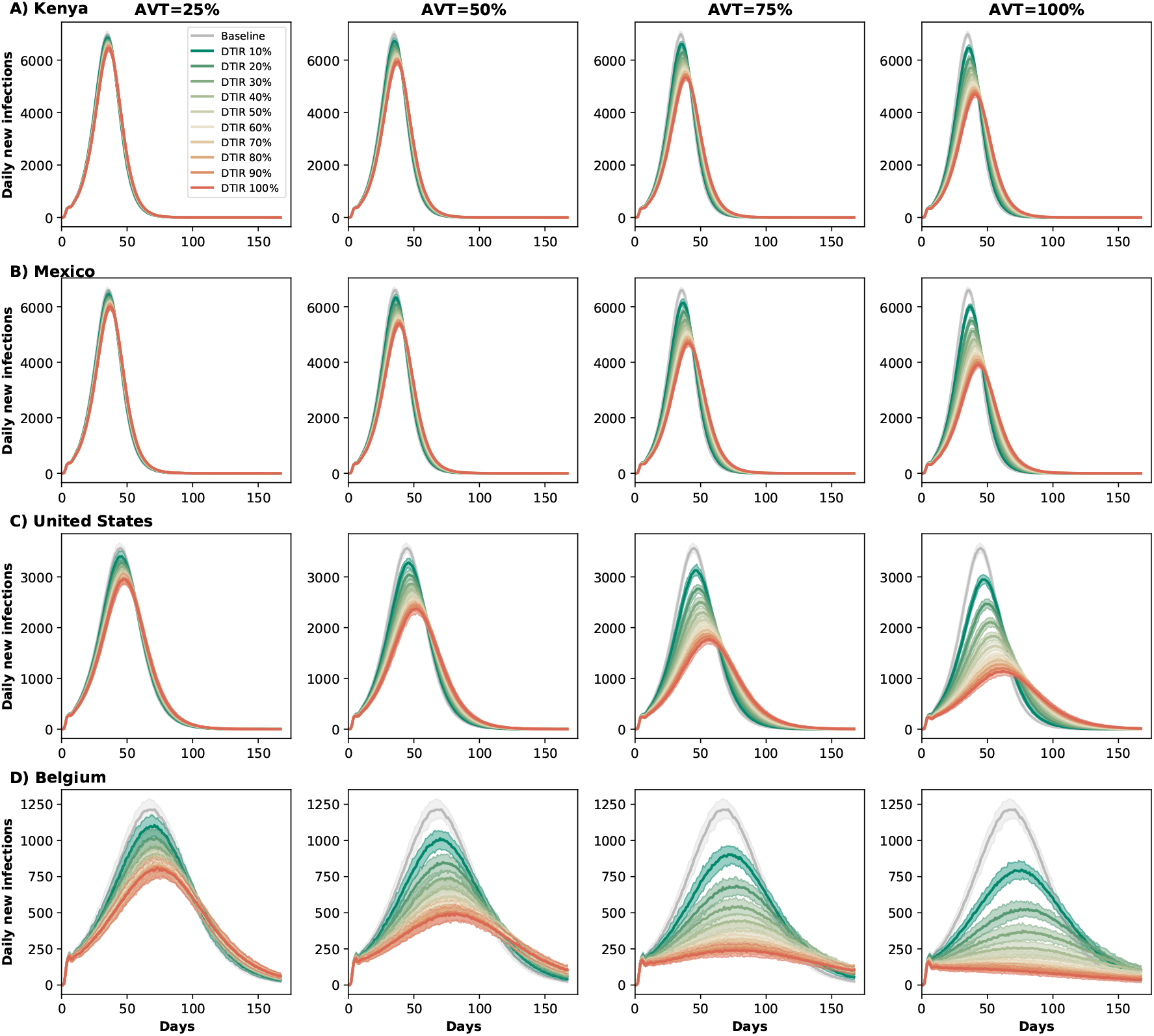
Daily new infections assuming no antiviral treatment (Baseline) or assuming a daily treatment initiation rate of of 10-100% of eligible symptomatic individuals in A) Kenya, B) Mexico, C) United States and D) Belgium assuming 60% of the population has been previously infected and is now recovered. For each location, each column represents a different value of AVT 25, 50, 75 or 100% reduction in viral transmission in treated symptomatic individuals).

**Figure S8:**
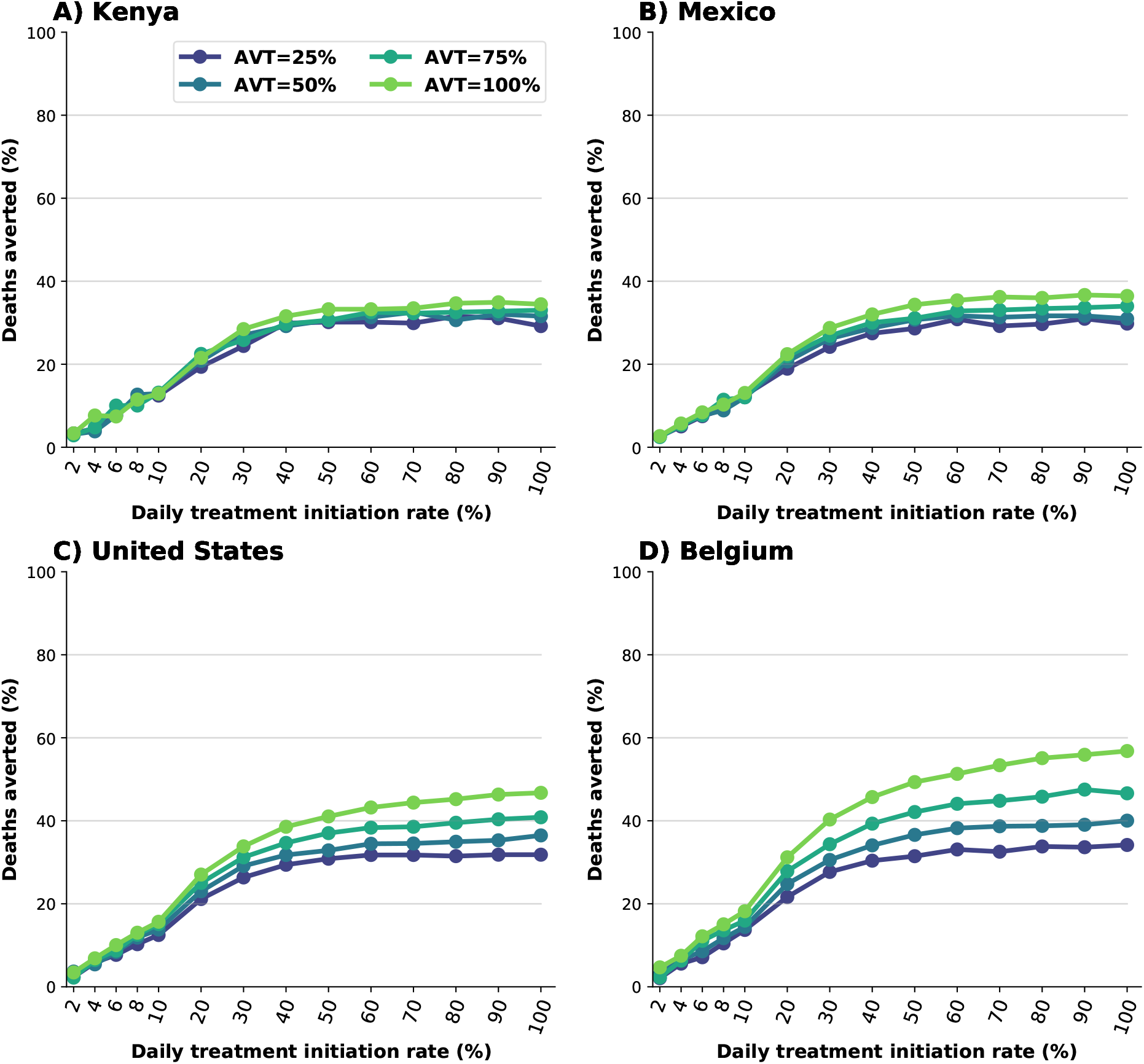
Percentage of deaths averted (compared to a baseline of no antiviral treatment) for A) Kenya, B) Mexico, C) United States and D) Belgium assuming antiviral treatment would reduce hospitalizations by 30%. Here, we assumed an epidemic wave with parameters similar to those of the Delta epidemic wave (transmissibility, vaccine effectiveness, and vaccination coverage). For each country, the colors represent four possible values of AVT (25, 50, 75 or 100% reduction in viral transmission in treated symptomatic individuals) and a daily treatment initiation rate (DTIR) of 2-100% of adult symptomatic individuals within the first 5 days of symptoms.

**Figure S9:**
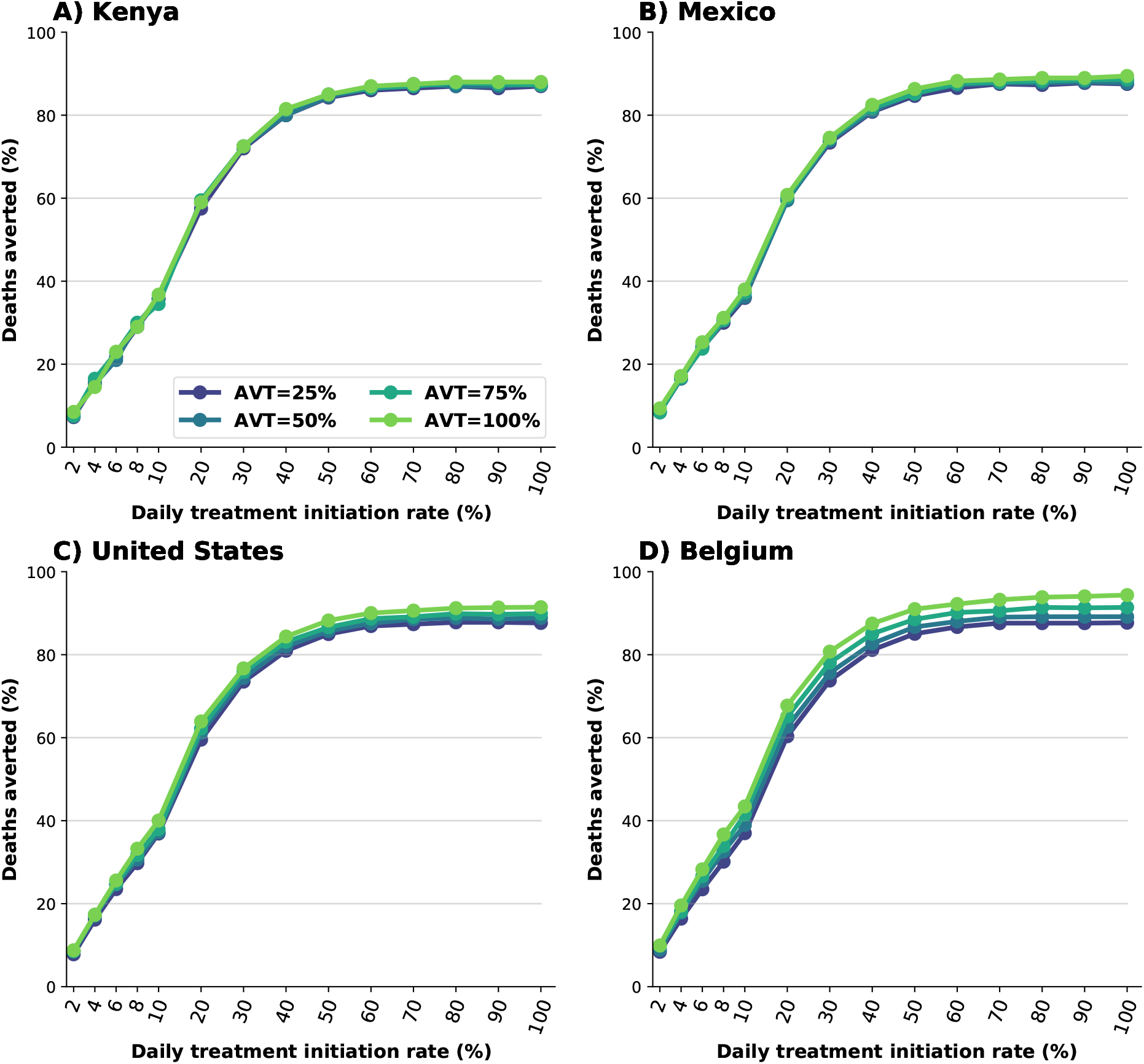
Percentage of deaths averted (compared to a baseline of no antiviral treatment) for A) Kenya, B) Mexico, C) United States and D) Belgium assuming asymptomatic infections are 50% less infectious. For each country, the colors represent four possible values of AVT (25, 50, 75 or 100% reduction in viral transmission in treated symptomatic individuals) and a daily treatment initiation rate (DTIR) of 2-100% of adult symptomatic individuals within the first 5 days of symptoms.

**Figure S10:**
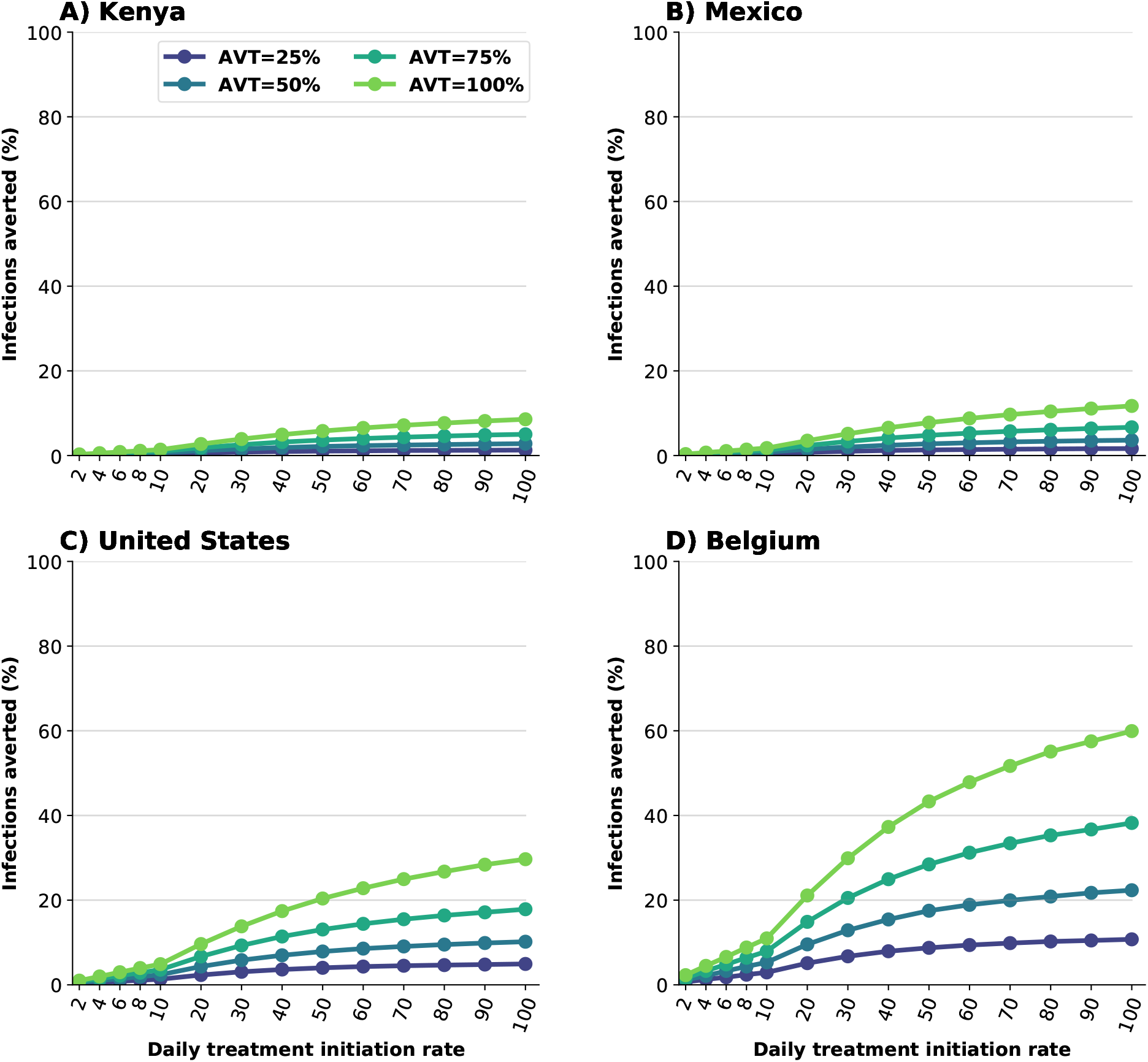
Percentage of infections averted (compared to a baseline of no antiviral treatment) for A) Kenya, B) Mexico, C) United States and D) Belgium assuming asymptomatic infections are 50% less infectious. For each country, the colors represent four possible values of AVT (25, 50, 75 or 100% reduction in viral transmission in treated symptomatic individuals) and a daily treatment initiation rate (DTIR) of 2-100% of adult symptomatic individuals within the first 5 days of symptoms.

## References

[1] Johns Hopkins Coronavirus Resource Center. Home - Johns Hopkins Coronavirus Resource Center. https://coronavirus.jhu.edu/, 2021. last accessed September 28th, 2021.

[2] Basta, Nicole and Moodie, Erika, on behalf of the McGill University COVID19 Vaccine Tracker Team. COVID-19 Vaccine Development and Approvals Tracker.. https://covid19.trackvaccines.org/, 2020. last accessed September 28th, 2021.

[3] Edouard Mathieu, Hannah Ritchie, Esteban Ortiz-Ospina, Max Roser, Joe Hasell, Cameron Appel, Charlie Giattino, and Lucas Rodés-Guirao. A global database of covid-19 vaccinations. Nature Human Behaviour, 5(7):947–953, 2021. doi: 10.1038/s41562-021-01122-8. URL https://doi.org/10.1038/s41562-021-01122-8.

[4] Jeffrey V. Lazarus, Scott C. Ratzan, Adam Palayew, Lawrence O. Gostin, Heidi J. Larson, Kenneth Rabin, Spencer Kimball, and Ayman El-Mohandes. A global survey of potential acceptance of a covid-19 vaccine. Nature Medicine, 27(2): 225–228, 2021. doi: 10.1038/s41591-020-1124-9. URL https://doi.org/10.1038/s41591-020-1124-9.

[5] Finlay Campbell, Brett Archer, Henry Laurenson-Schafer, Yuka Jinnai, Franck Konings, Neale Batra, Boris Pavlin, Katelijn Vandemaele, Maria D Van Kerkhove, Thibaut Jombart, Oliver Morgan, and Olivier le Polain de Waroux. Increased transmissibility and global spread of sars-cov-2 variants of concern as at june 2021. Eurosurveillance, 26(24):2100509, 2021. doi: https://doi.org/10.2807/1560-7917.ES.2021.26.24.2100509. URL https://www.eurosurveillance.org/content/10.2807/1560-7917.ES.2021.26.24.2100509.

[6] Kathryn Westendorf, Lingshu Wang, Stefanie Žentelis, Denisa Foster, Peter Vaillancourt, Matthew Wiggin, Erica Lovett, Robin van der Lee, Jörg Hendle, Anna Pustilnik, J. Michael Sauder, Lucas Kraft, Yuri Hwang, Robert W. Siegel, Jinbiao Chen, Beverly A. Heinz, Richard E. Higgs, Nicole Kallewaard, Kevin Jepson, Rodrigo Goya, Maia A. Smith, David W. Collins, Davide Pellacani, Ping Xiang, Valentine de Puyraimond, Marketa Ricicova, Lindsay Devorkin, Caitlin Pritchard, Aoise O’Neill, Kush Dalal, Pankaj Panwar, Harveer Dhupar, Fabian A. Garces, Courtney Cohen, John Dye, Kathleen E. Huie, Catherine V. Badger, Darwyn Kobasa, Jonathan Audet, Joshua J. Freitas, Saleema Hassanali, Ina Hughes, Luis Munoz, Holly C. Palma, Bharathi Ramamurthy, Robert W. Cross, Thomas W. Geisbert, Vineet Menacherry, Kumari Lokugamage, Viktoriya Borisevich, Iliana Lanz, Lisa Anderson, Payal Sipahimalani, Kizzmekia S. Corbett, Eun Sung Yang, Yi Zhang, Wei Shi, Tongqing Zhou, Misook Choe, John Misasi, Peter D. Kwong, Nancy J. Sullivan, Barney S. Graham, Tara L. Fernandez, Carl L. Hansen, Ester Falconer, John R. Mascola, Bryan E. Jones, and Bryan C. Barnhart. Ly-cov1404 (bebtelovimab) potently neutralizes sars-cov-2 variants. bioRxiv, 2022. doi: 10.1101/2021.04.30.442182. URL https://www.biorxiv.org/content/early/2022/01/07/ 2021.04.30.442182.

[7] Angélica Jayk Bernal, Monica M. Gomes da Silva, Dany B. Musungaie, Evgeniy Kovalchuk, Antonio Gonzalez, Virginia Delos Reyes, Alejandro Martín-Quirós, Yoseph Caraco, Angela Williams-Diaz, Michelle L. Brown, Jiejun Du, Alison Pedley, Christopher Assaid, Julie Strizki, Jay A. Grobler, Hala H. Shamsuddin, Robert Tipping, Hong Wan, Amanda Paschke, Joan R. Butterton, Matthew G. Johnson, and Carisa De Anda. Molnupiravir for oral treatment of covid-19 in nonhospitalized patients. New England Journal of Medicine, 386(6):509–520, 2022/02/15 2021. doi: 10.1056/NEJMoa2116044. URL https://doi.org/10.1056/NEJMoa2116044.

[8] Robert L. Gottlieb, Carlos E. Vaca, Roger Paredes, Jorge Mera, Brandon J. Webb, Gilberto Perez, Godson Oguchi, Pablo Ryan, Bibi U. Nielsen, Michael Brown, Ausberto Hidalgo, Yessica Sachdeva, Shilpi Mittal, Olayemi Osiyemi, Jacek Skarbinski, Kavita Juneja, Robert H. Hyland, Anu Osinusi, Shuguang Chen, Gregory Camus, Mazin Abdelghany, Santosh Davies, Nicole Behenna-Renton, Frank Duff, Francisco M. Marty, Morgan J. Katz, Adit A. Ginde, Samuel M. Brown, Joshua T. Schiffer, and Joshua A. Hill. Early remdesivir to prevent progression to severe covid-19 in outpatients. New England Journal of Medicine, 386(4):305–315, 2022/02/15 2021. doi: 10.1056/NEJMoa2116846. URL https://doi.org/10.1056/NEJMoa2116846.

[9] Anil Gupta, Yaneicy Gonzalez-Rojas, Erick Juarez, Manuel Crespo Casal, Jaynier Moya, Diego R. Falci, Elias Sarkis, Joel Solis, Hanzhe Zheng, Nicola Scott, Andrea L. Cathcart, Christy M. Hebner, Jennifer Sager, Erik Mogalian, Craig Tipple, Amanda Peppercorn, Elizabeth Alexander, Phillip S. Pang, Almena Free, Cynthia Brinson, Melissa Aldinger, and Adrienne E. Shapiro. Early treatment for covid-19 with sars-cov-2 neutralizing antibody sotrovimab. New England Journal of Medicine, 385(21):1941–1950, 2022/02/15 2021. doi: 10.1056/NEJMoa2107934. URL https://doi.org/10.1056/NEJMoa2107934.

[10] Pfizer. Pfizer’s Novel COVID-19 Oral Antiviral Treatment Candidate Reduced Risk of Hospitalization or Death by 89EPIC-HR Study | Pfizer. https://www.pfizer.com/news/press-release/press-release-detail/pfizers-novel-covid-19-oral-antiviral-treatment-candidate, Nov 2021. last accessed Nov 5th, 2021.

[11] FDA. Coronavirus Disease 2019 (COVID-19) EUA Information. https://www.fda.gov/emergency-preparedness-and-response/mcm-legal-regulatory-and-policy-framework/emergency-use-authorization#coviddrugs, February 2022. last accessed: February 28th, 2022.

[12] William Fischer, Joseph J Eron, Wayne Holman, Myron S Cohen, Lei Fang, Laura J Szewczyk, Timothy P Sheahan, Ralph Baric, Katie R Mollan, Cameron R Wolfe, Elizabeth R Duke, Masoud M Azizad, Katyna Borroto-Esoda, David A Wohl, Amy James Loftis, Paul Alabanza, Felicia Lipansky, and Wendy P Painter. Molnupiravir, an Oral Antiviral Treatment for COVID-19., Jun 2021.

[13] Pfizer. Pfizer Announces Additional Phase 2/3 Study Results Confirming Robust Efficacy of Novel COVID-19 Oral Antiviral Treatment Candidate in Reducing Risk of Hospitalization or Death. https://www.pfizer.com/news/press-release/press-release-detail/pfizer-announces-additional-phase-23-study-results, Dec 2021. last accessed March 18th, 2022.

[14] Centers for Disease Control and Prevention. The 2009 H1N1 Pandemic: Summary Highlights, April 2009-April 2010. https://www.cdc.gov/h1n1flu/cdcresponse.htm, mJune 2010. last accessed March 18th, 2022.

[15] Centers for Disease Control and Prevention. How to Optimize HIV Treatment | Treatment, Care, and Prevention for People with HIV | Clinicians | HIV | CDC. https://www.cdc.gov/hiv/clinicians/treatment/partner-prevention.html. last accessed October 12th, 2021.

[16] Myron S. Cohen, Ying Q. Chen, Marybeth McCauley, Theresa Gamble, Mina C. Hosseinipour, Nagalingeswaran Kumarasamy, James G. Hakim, Johnstone Kumwenda, Beatriz Grinsztejn, Jose H.S. Pilotto, Sheela V. Godbole, Suwat Chariyalertsak, Breno R. Santos, Kenneth H. Mayer, Irving F. Hoffman, Susan H. Eshleman, Estelle Piwowar-Manning, Leslie Cottle, Xinyi C. Zhang, Joseph Makhema, Lisa A. Mills, Ravindre Panchia, Sharlaa Faesen, Joseph Eron, Joel Gallant, Diane Havlir, Susan Swindells, Vanessa Elharrar, David Burns, Taha E. Taha, Karin Nielsen-Saines, David D. Celentano, Max Essex, Sarah E. Hudelson, Andrew D. Redd, and Thomas R. Fleming. Antiretroviral therapy for the prevention of hiv-1 transmission. New England Journal of Medicine, 375(9):830–839, 2016. doi: 10.1056/NEJMoa1600693. URL https://doi.org/10.1056/NEJMoa1600693. PMID: 27424812.

[17] Deborah Donnell, Jared M Baeten, James Kiarie, Katherine K Thomas, Wendy Stevens, Craig R Cohen, James McIntyre, Jairam R Lingappa, and Connie Celum. Heterosexual hiv-1 transmission after initiation of antiretroviral therapy: a prospective cohort analysis. The Lancet, 375(9731):2092–2098, 2010. ISSN 0140-6736. doi: https://doi.org/10.1016/S0140-6736(10)60705-2. URL https://www.sciencedirect.com/science/article/pii/S0140673610607052.

[18] FDA. Coronavirus (COVID-19) Update: FDA Authorizes Additional OTC Home Test to Increase Access to Rapid Testing for Consumers. https://www.fda.gov/news-events/press-announcements/coronavirus-covid-19-update-fda-authorizes-additional-otc-home-test-increase-access-rapid October 2021. last accessed March 18th, 2022.

[19] UNICEF. Most affordable COVID-19 rapid diagnostic test now available. https://www.unicef.org/supply/stories/most-affordable-covid-19-rapid-diagnostic-test-now-available, April 2021. last accessed March 18th, 2022.

[20] The White House. Remarks of President Joe Biden – State of the Union Address As Prepared for Delivery. https://www.whitehouse.gov/briefing-room/speeches-remarks/2022/03/01/remarks-of-president-joe-biden-state-of-the-union-address-as-delivered/, March 2022. last accessed March 11th, 2022.

[21] Medicines Patent Pool, UN. MPP and MSD announce new licence for investigational COVID-19 treatment. https://medicinespatentpool.org/news-publications-post/mpp-msd-new-licence-announcement-molnupiravir/, October 2021. URL https://medicinespatentpool.org/news-publications-post/mpp-msd-new-licence-announcement-molnupiravir/. last accessed October 27th, 2021.

[22] Pfizer. Pfizer and The Medicines Patent Pool (MPP) Sign Licensing Agreement for COVID-19 Oral Antiviral Treatment Candidate to Expand Access in Low-and Middle-Income Countries. https://www.pfizer.com/news/press-release/press-release-detail/pfizer-and-medicines-patent-pool-mpp-sign-licensing, Nov 2021. last accessed February 24, 2022.

[23] The New York Times. Pfizer will send 4 million courses of its Covid pill treatment to poorer countries. https://www.nytimes.com/live/2022/03/22/world/covid-19-mandates-cases-vaccine/pfizer-4-million-covid-pill-poorer-countries, March 2022. last accessed March 22, 2022.

[24] Cliff C. Kerr, Robyn M. Stuart, Dina Mistry, Romesh G. Abeysuriya, Katherine Rosenfeld, Gregory R. Hart, Rafael C. Núñez, Jamie A. Cohen, Prashanth Selvaraj, Brittany Hagedorn, Lauren George, Michal Jastrzębski, Amanda S. Izzo, Greer Fowler, Anna Palmer, Dominic Delport, Nick Scott, Sherrie L. Kelly, Caroline S. Bennette, Bradley G. Wagner, Stewart T. Chang, Assaf P. Oron, Edward A. Wenger, Jasmina Panovska-Griffiths, Michael Famulare, and Daniel J. Klein. Covasim: An agent-based model of covid-19 dynamics and interventions. PLOS Computational Biology, 17 (7):1–32, 07 2021. doi: 10.1371/journal.pcbi.1009149. URL https://doi.org/10.1371/journal.pcbi.1009149.

[25] Cliff C. Kerr, Dina Mistry, Robyn M. Stuart, Katherine Rosenfeld, Gregory R. Hart, Rafael C. Núñez, Jamie A. Cohen, Prashanth Selvaraj, Romesh G. Abeysuriya, Michal Jastrzębski, Lauren George, Brittany Hagedorn, Jasmina Panovska-Griffiths, Meaghan Fagalde, Jeffrey Duchin, Michael Famulare, and Daniel J. Klein. Controlling covid-19 via testtrace-quarantine. Nature Communications, 12(1):2993, 2021. doi: 10.1038/s41467-021-23276-9. URL https://doi.org/10.1038/s41467-021-23276-9.

[26] COVIDESTIM. covidestim: COVID-19 nowcasting. https://covidestim.org/, 2020. URL https://covidestim.org/. last accessed October 12th, 2021.

[27] Washington State Department of Health. COVID-19 Data Dashboard, 2021. URL https://www.doh.wa.gov/Emergencies/COVID19/DataDashboard{\#}dashboard. last accessed October 12th, 2021.

[28] Diana Buitrago-Garcia, Dianne Egli-Gany, Michel J. Counotte, Stefanie Hossmann, Hira Imeri, Aziz Mert Ipekci, Georgia Salanti, and Nicola Low. Occurrence and transmission potential of asymptomatic and presymptomatic sars-cov-2 infections: A living systematic review and meta-analysis. PLOS Medicine, 17(9):e1003346.#x2013;, 09 2020. URL https://doi.org/10.1371/journal.pmed.1003346.

[29] Hannah E. Davis, Gina S. Assaf, Lisa McCorkell, Hannah Wei, Ryan J. Low, Yochai Re’em, Signe Redfield, Jared P. Austin, and Athena Akrami. Characterizing long covid in an international cohort: 7 months of symptoms and their impact. eClinicalMedicine, 38, 2022/03/22 2021. doi: 10.1016/j.eclinm.2021.101019. URL https://doi.org/10.1016/j.eclinm.2021.101019.

[30] Maxime Taquet, Quentin Dercon, Sierra Luciano, John R. Geddes, Masud Husain, and Paul J. Harrison. Incidence, co-occurrence, and evolution of long-covid features: A 6-month retrospective cohort study of 273,618 survivors of covid-19. PLOS Medicine, 18(9):1–22, 09 2021. doi: 10.1371/journal.pmed.1003773. URL https://doi.org/10.1371/journal.pmed.1003773.

[31] Centers for Disease Control and Prevention. Post-COVID Conditions. https://www.cdc.gov/coronavirus/2019-ncov/long-term-effects/index.html, September 2021. last accessed March 18th, 2022.

[32] Xi He, Eric H.Y. Lau, Peng Wu, Xilong Deng, Jian Wang, Xinxin Hao, Yiu Chung Lau, Jessica Y. Wong, Yujuan Guan, Xinghua Tan, Xiaoneng Mo, Yanqing Chen, Baolin Liao, Weilie Chen, Fengyu Hu, Qing Zhang, Mingqiu Zhong, Yanrong Wu, Lingzhai Zhao, Fuchun Zhang, Benjamin J. Cowling, Fang Li, and Gabriel M. Leung. Temporal dynamics in viral shedding and transmissibility of COVID-19. Nature Medicine, 26(May), 2020. ISSN 1546170X. doi: 10.1038/s41591-020-0869-5.

[33] Ben Killingley, Alex Mann, Mariya Kalinova, Alison Boyers, Niluka Goonawardane, Jie Zhou, Kate Lindsell, Samanjit S. Hare, Jonathan Brown, Rebecca Frise, Emma Smith, Claire Hopkins, Nicolas Noulin, Brandon Londt, Tom Wilkinson, Stephen Harden, Helen McShane, Mark Baillet, Anthony Gilbert, Michael Jacobs, Christine Charman, Priya Mande, Jonathan S. Nguyen-Van-Tam, Malcolm G. Semple, Robert C. Read, Neil M. Ferguson, Peter J. Openshaw, Garth Rapeport, Wendy S. Barclay, Andrew P. Catchpole, and Christopher Chiu. Safety, tolerability and viral kinetics during SARS-CoV-2 human challenge. Nature Portfolio, 2022. doi: 10.21203/rs.3.rs-1121993/v1. URL https://doi.org/10.21203/rs.3.rs-1121993/v1.

[34] Youngji Jo, Lise Jamieson, Ijeoma Edoka, Lawrence Long, Sheetal Silal, Juliet R.C. Pulliam, Harry Moultrie, Ian Sanne, Gesine Meyer-Rath, and Brooke E Nichols. Cost-effectiveness of remdesivir and dexamethasone for covid-19 treatment in south africa. medRxiv, 2020. doi: 10.1101/2020.09.24.20200196. URL https://www.medrxiv.org/content/early/2020/09/27/2020.09.24.20200196.

[35] Ashish Goyal, E. Fabian Cardozo-Ojeda, and Joshua T. Schiffer. Potency and timing of antiviral therapy as determinants of duration of sars-cov-2 shedding and intensity of inflammatory response. Science Advances, 6(47):eabc7112, 2021/11/08 2020. doi: 10.1126/sciadv.abc7112. URL https://doi.org/10.1126/sciadv.abc7112.

[36] Kathy Leung, Mark Jit, Gabriel M Leung, and Joseph T Wu. Comparative effectiveness of allocation strategies of covid-19 vaccines and antivirals against emerging sars-cov-2 variants of concern in east asia and pacific region. medRxiv, 2021. doi: 10.1101/2021.10.20.21265245. URL https://www.medrxiv.org/content/early/2021/10/20/2021.10.20.21265245.

[37] Andrea Torneri, Pieter Libin, Joris Vanderlocht, Anne-Mieke Vandamme, Johan Neyts, and Niel Hens. A prospect on the use of antiviral drugs to control local outbreaks of covid-19. medRxiv, 2020. doi: 10.1101/2020.03.19.20038182. URL https://www.medrxiv.org/content/early/2020/03/30/2020.03.19.20038182.

[38] Charles Whittaker, Oliver J. Watson, Carlos Alvarez-Moreno, Nasikarn Angkasekwinai, Adhiratha Boonyasiri, Luis Carlos Triana, Duncan Chanda, Lantharita Charoenpong, Methee Chayakulkeeree, Graham S. Cooke, Julio Croda, Zulma M Cucunubá, Bimandra A. Djaafara, Cassia F. Estofolete, Maria Eugenia Grillet, Nuno R. Faria, Silvia Figueiredo Costa, David A. Forero-Peña, Diana M. Gibb, Anthony C Gordon, Raph L. Hamers, Arran Hamlet, Vera Irawany, Anupop Jitmuang, Nukool Keurueangkul, Teresia Njoki Kimani, Margarita Lampo, Anna S. Levin, Gustavo Lopardo, Rima Mustafa, Shevanthi Nayagam, Thundon Ngamprasertchai, Ng’ang’a Irene Hannah Njeri, Mauricio L. Nogueira, Esteban Ortiz-Prado, Mauricio W. Perroud, Andrew N. Phillips, Panuwat Promsin, Ambar Qavi, Alison J. Rodger, Ester C. Sabino, Sorawat Sangkaew, Djayanti Sari, Rujipas Sirijatuphat, Andrei C. Sposito, Pratthana Srisangthong, Hayley A. Thompson, Zarir Udwadia, Sandra Valderrama-Beltrán, Peter Winskill, Azra C. Ghani, Patrick G.T. Walker, and Timothy B. Hallett. Understanding the potential impact of different drug properties on sars-cov-2 transmission and disease burden: A modelling analysis. medRxiv, 2021. doi: 10.1101/2021.06.17.21259078. URL https://www.medrxiv.org/content/early/2021/06/20/2021.06.17.21259078.

[39] Anupriya Aggarwal, Alberto Ospina Stella, Gregory Walker, Anouschka Akerman, Vanessa Milogiannakis, Fabienne Brilot, Supavadee Amatayakul-Chantler, Nathan Roth, Germano Coppola, Peter Schofield, Jennifer Jackson, Jake Y. Henry, Ohan Mazigi, David Langley, Yonghui Lu, Charles Forster, Samantha McAllery, Vennila Mathivanan, Christina Fichter, Alexandra Carey Hoppe, Mee Ling Munier, Hans-Martin Jack, Deborah Cromer, David Darley, Gail Matthews, Daniel Christ, David Khoury, Miles Davenport, William Rawlinson, Anthony D. Kelleher, and Stuart Turville. Sars-cov-2 omicron: evasion of potent humoral responses and resistance to clinical immunotherapeutics relative to viral variants of concern. medRxiv, 2021. doi: 10.1101/2021.12.14.21267772. URL https://www.medrxiv.org/content/early/2021/12/15/2021.12.14.21267772.

[40] Hao Zhou, Takuya Tada, Belinda M. Dcosta, and Nathaniel R. Landau. Sars-cov-2 omicron ba.2 variant evades neutralization by therapeutic monoclonal antibodies. bioRxiv, 2022. doi: 10.1101/2022.02.15.480166. URL https://www.biorxiv.org/content/early/2022/02/16/2022.02.15.480166.

[41] Emi Takashita, Noriko Kinoshita, Seiya Yamayoshi, Yuko Sakai-Tagawa, Seiichiro Fujisaki, Mutsumi Ito, Kiyoko Iwatsuki-Horimoto, Peter Halfmann, Shinji Watanabe, Kenji Maeda, Masaki Imai, Hiroaki Mitsuya, Norio Ohmagari, Makoto Takeda, Hideki Hasegawa, and Yoshihiro Kawaoka. Efficacy of Antiviral Agents against the SARS-CoV-2 Omicron Subvariant BA.2. https://doi.org/10.1056/NEJMc2201933, March 2022. URL https://doi.org/10.1056/NEJMc2201933. last accessed March 18th, 2022.

[42] CM Brown, J Vostok, H Johnson, and et al. Outbreak of SARS-CoV-2 Infections, Including COVID-19 Vaccine Break-through Infections, Associated with Large Public Gatherings — Barnstable County, Massachusetts, July 2021. MMWR Morb Mortal Wkly Rep, 70:1059–1062, August 2021.

[43] James A. Hay, Stephen M. Kissler, Joseph R. Fauver, Christina Mack, Caroline G. Tai, Radhika M. Samant, Sarah Connelly, Deverick J. Anderson, Gaurav Khullar, Matthew MacKay, Miral Patel, Shannan Kelly, April Manhertz, Isaac Eiter, Daisy Salgado, Tim Baker, Ben Howard, Joel T. Dudley, Christopher E. Mason, David D. Ho, Nathan D. Grubaugh, and Yonatan H. Grad. Viral dynamics and duration of pcr positivity of the sars-cov-2 omicron variant. medRxiv, 2022. doi: 10.1101/2022.01.13.22269257. URL https://www.medrxiv.org/content/early/2022/01/14/2022.01.13.22269257.

[44] Maylis Layan, Mayan Gilboa, Tal Gonen, Miki Goldenfeld, Lilac Meltzer, Alessio Andronico, Nathanaël Hozé, Simon Cauchemez, and Gili Regev-Yochay. Impact of BNT162b2 vaccination and isolation on SARS-CoV-2 transmission in Israeli households: an observational study. American Journal of Epidemiology, 03 2022. ISSN 0002-9262. doi: 10.1093/aje/kwac042. URL https://doi.org/10.1093/aje/kwac042. kwac042.

[45] Ottavia Prunas, Joshua L. Warren, Forrest W. Crawford, Sivan Gazit, Tal Patalon, Daniel M. Weinberger, and Virginia E. Pitzer. Vaccination with bnt162b2 reduces transmission of sars-cov-2 to household contacts in israel. Science, 375 (6585):1151–1154, 2022. doi: 10.1126/science.abl4292. URL https://www.science.org/doi/abs/10.1126/science.abl4292.

[46] WHO. WHO, UN set out steps to meet world COVID vaccination targets. https://www.who.int/news/item/07-10-2021-who-un-set-out-steps-to-meet-world-covid-vaccination-targets, 2021. URL https://www.who.int/news/item/07-10-2021-who-un-set-out-steps-to-meet-world-covid-vaccination

[47] Hannah W Despres, Margaret G Mills, David J Shirley, Madaline M Schmidt, Meei-Li Huang, Keith R Jerome, Alexander L Greninger, and Emily A Bruce. Quantitative measurement of infectious virus in sars-cov-2 alpha, delta and epsilon variants reveals higher infectivity (viral titer:rna ratio) in clinical samples containing the delta and epsilon variants. medRxiv, Sep 2021. doi: 10.1101/2021.09.07.21263229.

[48] Q Bi, J Lessler, I Eckerle, S A Lauer, L Kaiser, N Vuilleumier, D A T Cummings, A Flahault, D Petrovic, I Guessous, S Stringhini, A S Azman, and SeroCoV-POP Study Group. Household transmission of SARS-CoV-2: Insights from a population-based serological survey. medRxiv, 2021. URL https://www.medrxiv.org/content/10.1101/2020.11.04.20225573v2.

[49] Christelle Meuris, Cécile Kremer, Anton Geerinck, Medea Locquet, Olivier Bruyère, Justine Defêche, Cécile Meex, Marie-Pierre Hayette, Loic Duchene, Patricia Dellot, Samira Azarzar, Nicole Maréchal, Anne-Sophie Sauvage, Frederic Frippiat, Jean-Baptiste Giot, Philippe Léonard, Karine Fombellida, Michel Moutschen, Keith Durkin, Maria Artesi, Vincent Bours, Christel Faes, Niel Hens, and Gilles Darcis. Transmission of SARS-CoV-2 After COVID-19 Screening and Mitigation Measures for Primary School Children Attending School in Liège, Belgium. JAMA Network Open, 4 (10):e2128757–e2128757, 10 2021. ISSN 2574-3805. doi: 10.1001/jamanetworkopen.2021.28757. URL https://doi.org/10.1001/jamanetworkopen.2021.28757.

[50] CDC. COVID-19 Pandemic Planning Scenarios. URL https://www.cdc.gov/coronavirus/2019-ncov/hcp/planning-scenarios.html.

[51] Carl A. B. Pearson, Sheetal P. Silal, Michael W.Z. Li, Jonathan Dushoff, Benjamin M. Bolker, Sam Abbott, Cari van Schalkwyk, Nicholas G. Davies, Rosanna C. Barnard, W. John Edmunds, Jeremy Bingham, Gesine Meyer-Rath, Lise Jamieson, Allison Glass, Nicole Wolter, Nevashan Govender, Wendy S. Stevens, Lesley Scott, Koleka Mlisana, Harry Moultrie, and Juliet R. C. Pulliam. Bounding the levels of transmissibility & immune evasion of the omicron variant in south africa. medRxiv, 2021. doi: 10.1101/2021.12.19.21268038. URL https://www.medrxiv.org/content/early/2021/12/21/2021.12.19.21268038.

[52] M Elizabeth Halloran, Claudio J Struchiner, and Ira M Longini. Study designs for evaluating different efficacy and effectiveness aspects of vaccines. American Journal of Epidemiology, 146(10):789–803, 1997. ISSN 0002-9262 (Print).

[53] Secretaria de Salud, Mexico. Vacuna COVID. http://vacunacovid.gob.mx/wordpress/, 2021. last accessed October 12th, 2021.

[54] Centers for Disease Control and Prevention. COVID-19 Vaccination and Case Trends by Age Group, United States Vaccinations, 2021. URL https://data.cdc.gov/Vaccinations/COVID-19-Vaccination-and-Case-Trends-by-Age-Group-/gxj9-t96f. last accessed October 12th, 2021.

[55] Sciensano. Belgium COVID-19 Dashboard - Sciensano › Vaccination, 2021. URL https://datastudio. google.com/u/0/reporting/c14a5cfc-cab7-4812-848c-0369173148ab/page/hOMwB. last accessed October 12th, 2021.

[56] http://OurWorldInData.org. COVID-19 vaccine doses, people with at least one dose, people fully vaccinated, and boost- ers per 100 people. https://ourworldindata.org/explorers/coronavirus-data-explorer?zoomToSelection=true&time=2020-03-01..latest&uniformYAxis=0&pickerSort=asc&pickerMetric=location&Metric=Vaccine+doses%2C+people+vaccinated%2C+and+booster+doses&Interval=7-day+rolling+average&Relative+to+Population=false&Color+by+test+positivity=false&country=USA∼GBR∼CAN∼DEU∼ITA∼IND,2021. last accessed February 24, 2022.

[57] GAVI. Kenya completes its first round of COVID-19 vaccinations | Gavi, the Vaccine Alliance. https://www.gavi.org/vaccineswork/kenya-completes-its-first-round-covid-19-vaccinations, 2021. last accessed October 12th, 2021.

[58] Stephen A Lauer, Kyra H Grantz, Qifang Bi, Forrest K Jones, Qulu Zheng, Hannah R Meredith, Andrew S Azman, Nicholas G Reich, and Justin Lessler. The incubation period of coronavirus disease 2019 (COVID-19) from publicly reported confirmed cases: Estimation and application. Annals of Internal Medicine, 2020. ISSN 1539-3704. doi: 10.7326/M20-0504. URL http://www.ncbi.nlm.nih.gov/pubmed/32150748.

[59] Roman Wölfel, Victor M. Corman, Wolfgang Guggemos, Michael Seilmaier, Sabine Zange, Marcel A. Müller, Daniela Niemeyer, Terry C. Jones, Patrick Vollmar, Camilla Rothe, Michael Hoelscher, Tobias Bleicker, Sebastian Brünink, Julia Schneider, Rosina Ehmann, Katrin Zwirglmaier, Christian Drosten, and Clemens Wendtner. Virological assessment of hospitalized patients with covid-2019. Nature, 581(7809):465–469, 2020. doi: 10.1038/s41586-020-2196-x. URL https://doi.org/10.1038/s41586-020-2196-x.

[60] Natalie M Linton, Tetsuro Kobayashi, Yichi Yang, Katsuma Hayashi, Andrei R Akhmetzhanov, Sung-Mok Jung, Baoyin Yuan, Ryo Kinoshita, and Hiroshi Nishiura. Incubation period and other epidemiological characteristics of 2019 novel coronavirus infections with right truncation: A statistical analysis of publicly available case data. Journal of clinical medicine, 9(2):538, 02 2020. doi: 10.3390/jcm9020538. URL https://pubmed.ncbi.nlm.nih.gov/32079150.

[61] Dawei Wang, Bo Hu, Chang Hu, Fangfang Zhu, Xing Liu, Jing Zhang, Binbin Wang, Hui Xiang, Zhenshun Cheng, Yong Xiong, Yan Zhao, Yirong Li, Xinghuan Wang, and Zhiyong Peng. Clinical Characteristics of 138 Hospitalized Patients With 2019 Novel Coronavirus–Infected Pneumonia in Wuhan, China. JAMA, 323(11):1061–1069, 03 2020. ISSN 0098-7484. doi: 10.1001/jama.2020.1585. URL https://doi.org/10.1001/jama.2020.1585.

[62] Jun Chen, Tangkai Qi, Li Liu, Yun Ling, Zhiping Qian, Tao Li, Feng Li, Qingnian Xu, Yuyi Zhang, Shuibao Xu, Zhigang Song, Yigang Zeng, Yinzhong Shen, Yuxin Shi, Tongyu Zhu, and Hongzhou Lu. Clinical progression of patients with covid-19 in shanghai, china. Journal of Infection, 80(5):e1–e6, 2020. ISSN 0163-4453. doi: https://doi.org/10.1016/j.jinf.2020.03.004. URL https://www.sciencedirect.com/science/article/pii/S0163445320301195.

[63] Robert Verity, Lucy C Okell, Ilaria Dorigatti, Peter Winskill, Charles Whittaker, Natsuko Imai, Gina Cuomo-Dannenburg, Hayley Thompson, Patrick G T Walker, Han Fu, Amy Dighe, Jamie T Griffin, Marc Baguelin, Sangeeta Bhatia, Adhiratha Boonyasiri, Anne Cori, Zulma Cucunubá, Rich FitzJohn, Katy Gaythorpe, Will Green, Arran Hamlet, Wes Hinsley, Daniel Laydon, Gemma Nedjati-Gilani, Steven Riley, Sabine van Elsland, Erik Volz, Haowei Wang, Yuanrong Wang, Xiaoyue Xi, Christl A Donnelly, Azra C Ghani, and Neil M Ferguson. Estimates of the severity of coronavirus disease 2019: a model-based analysis. The Lancet Infectious Diseases, 20(6):669–677, 2021/11/01 2020. doi: 10.1016/S1473-3099(20)30243-7. URL https://doi.org/10.1016/S1473-3099(20)30243-7.

[64] Megan O’Driscoll, Gabriel Ribeiro Dos Santos, Lin Wang, Derek A. T. Cummings, Andrew S. Azman, Juliette Paireau, Arnaud Fontanet, Simon Cauchemez, and Henrik Salje. Age-specific mortality and immunity patterns of sars-cov-2. Nature, 590(7844):140–145, 2021. doi: 10.1038/s41586-020-2918-0. URL https://doi.org/10.1038/s41586-020-2918-0.

[65] Nick F Brazeau, Robert Verity, Sara Jenks, Han Fu, Charles Whittaker, Pete Winskill, Ilaria Dorigatti, Patrick G T Walker, Steven Riley, Ricardo P Schnekenbert, Henrique Hoeltgebaum, Thomas A Mellan, Swapnil Mishra, H Juliette T Unwin, Oliver J Watson, Zulma Cucunuba, Marc Baguelin, Lilith K Whittles, Samir Bhatt, Azra C. Ghani, Ferguson Neil M., and Lucy C Okell. Report 34 - COVID-19 Infection Fatality Ratio Estimates from Seroprevalence. https://www.imperial.ac.uk/mrc-global-infectious-disease-analysis/covid-19/report-34-ifr/, 2020.

[66] Neil M Ferguson, Daniel Laydon, Gemma Nedjati-Gilani, Natsuko Imai, Kylie Ainslie, Marc Baguelin, Sangeeta Bhatia, Adhiratha Boonyasiri, Zulma Cucunubá, Gina Cuomo-Dannenburg, Amy Dighe, Ilaria Dorigatti, Han Fu, Katy Gaythorpe, Will Green, Arran Hamlet, Wes Hinsley, Lucy C Okell, Sabine van Elsland, Hayley Thompson, Robert Verity, Erik Volz, Haowei Wang, Yuanrong Wang, Patrick G T Walker, Caroline Walters, Peter Winskill, Charles Whittaker, Christl A Donnelly, Steven Riley, and Azra C Ghani. Impact of non-pharmaceutical interventions (NPIs) to reduce COVID-19 mortality and healthcare demand. 2020. URL https://doi.org/10.25561/77482.

[67] Russell M. Viner, Oliver T. Mytton, Chris Bonell, G. J. Melendez-Torres, Joseph Ward, Lee Hudson, Claire Waddington, James Thomas, Simon Russell, Fiona Van Der Klis, Archana Koirala, Shamez Ladhani, Jasmina Panovska-Griffiths, Nicholas G. Davies, Robert Booy, and Rosalind M. Eggo. Susceptibility to SARS-CoV-2 infection among children and adolescents compared with adults: A systematic review and meta-analysis. JAMA Pediatrics, 175(2):143–156, 2021. ISSN 21686211. doi: 10.1001/jamapediatrics.2020.4573.

[68] Yong-Hoon Lee, Chae Moon Hong, Dae Hyun Kim, Taek Hoo Lee, and Jaetae Lee. Clinical course of asymptomatic and mildly symptomatic patients with coronavirus disease admitted to community treatment centers, South Korea. Emerging Infectious Diseases, Oct, 2020. doi: 10.3201/eid2610.201620. URL https://www.nc.cdc.gov/eid/article/26/10/20-1620{\_}article.

[69] Kenji Mizumoto, Katsushi Kagaya, Alexander Zarebski, and Gerardo Chowell. Estimating the asymptomatic proportion of coronavirus disease 2019 (COVID-19) cases on board the Diamond Princess cruise ship, Yokohama, Japan, 2020. Euro Surveillance, 25(10), 2020. URL doi.org/10.2807/1560-7917.

[70] Gilead. Veklury® (Remdesivir) Significantly Reduced Risk of Hospitalization in High-Risk Patients with COVID-19. https://www.gilead.com/news-and-press/press-room/press-releases/2021/9/veklury-remdesivir-significantly-reduced-risk-of-hospitalization-in-highrisk-patients-with-September 2021. last accessed October 19th, 2021.

[71] Merck. Merck and Ridgeback’s Investigational Oral Antiviral Molnupiravir Reduced the Risk of Hospitalization or Death by Approximately 50 Percent Compared to Placebo for Patients with Mild or Moderate COVID-19 in Positive Interim Analysis of Phase 3 Study - Merc. https://www.merck.com/news/merck-and-ridgebacks-investigational-oral-antiviral-molnupiravir-reduced-the-risk-of-hospitalization October 2021. last accessed October 19th, 2021.

[72] Centers for Disease Control and Prevention. CDC COVID Data Tracker. https://covid.cdc.gov/covid-data-tracker/\#vaccine-effectiveness, 2020. last accessed October 19th, 2021.

[73] Neil Ferguson, Azra Ghani, Anne Cori, Alexandra Hogan, Wes Hinsley, Erik Volz, and on behalf of the Imperial College COVID-19 response Team. Report 49 - Growth, population distribution and immune escape of Omicron in England | Faculty of Medicine | Imperial College London, 2021. URL https://www.imperial.ac.uk/mrc-global-infectious-disease-analysis/covid-19/report-49-Omicron/.

[74] UK Health Security Agency. SARS-CoV-2 variants of concern and variants under investigation in England, Technical Briefing 31. https://assets.publishing.service.gov.uk/government/uploads/system/uploads/attachment\_data/file/1042367/technical\_briefing-31-10-december-2021.pdf, Dec 2021. last accessed February 24, 2022.

